# Burden of Disease of Respiratory Syncytial Virus in Older Adults and Adults Considered at High Risk of Severe Infection

**DOI:** 10.1101/2024.03.18.24304476

**Authors:** Elissa Abrams, Pamela Doyon-Plourde, Phaedra Davis, Liza Lee, Abbas Rahal, Nicholas Brousseau, Winnie Siu, April Killikelly

## Abstract

**Background:** Vaccine products for adults have increased interest in understanding Canada’s respiratory syncytial virus (RSV) burden in older adults and adults considered at high risk of severe infection.

**Objective:** To characterize the burden of RSV disease in Canada by joint analysis of the published literature and hospitalization data from a healthcare administrative database.

**Methods:** Electronic databases of published literature were searched to identify studies and systematic reviews reporting data on outpatient visits, hospitalizations, intensive care unit (ICU) admissions and deaths associated with RSV infection in adults. For the hospitalization data analysis, hospital discharge records were extracted from the Canadian Institute of Health Information (CIHI) Discharge Abstract Database (DAD) for all patients admitted to an acute care facility for RSV infection defined by ICD-10 codes from 2010-2020 and 2021-2023.

**Results:** Overall, 26 studies, including 7 systematic reviews, were identified and summarized in the rapid review. Evidence suggests that medically attended RSV respiratory tract infections (RTI) are frequent causing 4.7 to 7.8% of symptomatic RTI in adults 60 years of age and older. Incidence of RSV RTI increases with age and presence of underlying medical conditions, such as cardiorespiratory disease, diabetes, and immunocompromising conditions. This trend was consistently observed across all RSV clinical outcomes of interest (i.e., hospitalization, ICU admission and death). Patients who reside in long-term care or other chronic care facilities have higher likelihood of severe clinical outcomes (i.e., ICU admission, receiving mechanical ventilation and/or death) compared to patients with other living situations upon hospital admission. Approximately 10% of older adults hospitalized with RSV infection require ICU admission. Although data are limited, evidence suggests that case fatality ratio (CFR) among those admitted to hospital varies between 5 and 10%. Some evidence suggests that RSV burden may be close to influenza burden in older adults. In general, the results from the Canadian hospitalization data support the rapid review findings Rates of hospitalization, ICU admission and death associated with RSV all increased with age, with 16% of hospitalizations resulting in ICU admission and with an in-hospital CFR of 9%.

**Conclusion:** In adults, risk of severe RSV outcomes in general increases with increasing age and presence of comorbidities.

## Introduction

Respiratory syncytial virus (RSV) is commonly recognized as a significant respiratory pathogen mostly affecting young children under 24 months of age, and older adults. Although the burden of disease in the older adult demographic can be substantial, with older adults experiencing more severe disease compared to younger populations, this is not as well described as it is in children and for other pathogens. It has been estimated that globally, RSV is associated with approximately 336,000 hospitalizations and 14,000 in-hospital deaths each year in adults 65 years and older (1). Additionally, evidence suggest that younger adults living with underlying medical conditions, such as immunocompromising conditions and chronic cardiopulmonary disease, are at high risk of severe RSV infection and complications (2, 3). Nonetheless, RSV remains generally underrecognized as a cause of severe respiratory tract infection (RTI) in adults.

The RSV vaccine landscape has evolved dramatically for older adults and potentially younger adults with certain risk factors in the past year. While previously there were no vaccine products available, there are currently three RSV vaccines being considered in Canada. As of February 2024, The GSK RSVPreF3 vaccine (Arexvy) and the Pfizer RSVpreF vaccine (Abrysvo) have been approved by Health Canada, and the Moderna m1345 RSV vaccine is being reviewed by Health Canada. As vaccination will be available to older adults for the first time, there is a need for a more granular understanding of the burden of RSV disease to inform vaccine recommendations. Therefore, this rapid review aimed to evaluate RSV burden of disease in adults from high income countries (Canada, United States, European countries, Australia). Additionally, hospitalization data from the Canadian Institute for Health Information (CIHI) Discharge Abstract Database (DAD) were analyzed to further describe RSV burden in Canada. This report compiles evidence derived from the literature and the Canadian administrative health database in order to present a comprehensive picture of RSV burden of disease to inform immunization guidance development in adults.

## Methods

### Rapid Review

#### Search Strategies

The search strategy was developed by a research librarian from Health Canada and the Public Health Agency of Canada. EMBASE, MEDLINE, Global Heath and ProQuest Public Health databases were searched from 1995 to November 2022, and again on September 1^st^ 2023, to identify recent studies evaluating RSV burden of disease in adults (Supplementary material S1-S6). Canadian respiratory virus surveillance experts were also contacted for any additional data. After removal of duplicates, references were uploaded in DistillerSR online software (Evidence Partners Inc, Ottawa, Ontario).

#### Study Selection

Two reviewers screened titles and abstracts for study eligibility. Full texts of selected studies were then evaluated. A second independent reviewer assessed citations marked for exclusion, with disagreements resolved through discussion. Reference lists of included studies were also screened for relevant articles on RSV burden in high-income countries.

#### Eligibility Criteria

Inclusion was limited to studies reporting data on RSV infection in adults, with a focus on adults 50 years of age and older and individuals 18 years of age and older with underlying medical conditions. The evaluation of RSV burden of disease focused on clinical outcomes of interest including medically attended RSV RTI, hospitalizations, intensive care unit (ICU) admissions, and death associated with RSV infection (Supplementary material S7). Observational studies, randomized controlled trials (RCTs) and systematic reviews (SRs) were included. Exclusion criteria were populations of other ages, and studies that did not reported on outcomes of interest.

#### Data Extraction and Data Synthesis

One reviewer extracted data from each article, verified by a second reviewer. Disagreements were resolved through discussion. Data extracted included study design, study period, population characteristics, outcome definitions, sample size, number of events and effect measures. When reported in included studies, results comparing RSV and influenza burden of disease were extracted. Results were synthesized narratively based on the study population and outcomes. Subgroups of interest included long-term care (LTC) residents, adults with immunocompromising conditions, and adults with chronic medical conditions.

### Canadian Hospitalization Data

#### Data Source

Hospital discharge records were extracted from the CIHI DAD which contains data from acute care facilities from all provinces and territories, except Quebec, representing 78% of the Canadian population (4). Population demographic data (i.e., age group) were extracted from the Statistics Canada website (5).

#### Analytic Cohort

All patients admitted to an acute care facility with RSV between September 2010 to August 2020 and September 2021 to August 2023 (12 respiratory virus seasons spanning September through August of the following year) were included in the analysis. Due to public health measures enacted for the coronavirus disease 2019 (COVID-19) pandemic, there was almost no RSV activity in the 2020-2021 season (6); therefore, this season was excluded from the analysis as it did not reflect normal seasonal activity.

Hospitalization associated with RSV was identified using the International Classification of Disease, Tenth Modification (ICD-10) codes J12.1, J20.5, J21.0 or B97.4 found anywhere from diagnosis 1 through 25. Hospitalization due to RSV was defined as one of aforementioned ICD-10 codes recorded as diagnosis 1. Data on ICU admissions and in-hospital deaths were also extracted. Diagnosis codes were not available for ICU admissions and deaths; therefore, their classifications (associated with or due to RSV) were based on whether the initial hospitalization was associated with or due to RSV.

Risk factors of interest were also determined by diagnosis information based on ICD-10 classification codes and were chosen based on prior known associations with severe RSV outcomes. Diagnosis codes were considered mutually exclusive (i.e. one individual hospitalized for RSV with multiple risk factors of interest were counted in each individual risk factor category). All diagnoses and conditions that are present on a patient’s record from diagnosis 1 through 25 were included in determining their risk factors. Risk factors of interest for the Canadian hospitalization data analysis included RTI, chronic obstructive pulmonary disease (COPD), immunocompromising conditions, cardiovascular disease, diabetes and chronic kidney disease. The list of ICD-10 codes used to define a risk factor is found in Supplementary material (S8).

#### Data Synthesis

The number of hospitalizations, ICU admissions and in-hospital death analyses were both aggregated and stratified by season and age groups where appropriate. Hospitalizations were also presented as rates aggregated by season and stratified by age groups (50-59 years, 60-69 years, 70-79 years and ≥80 years). Moreover, ICU admission rates and case fatality ratios (CFR) were presented by age group aggregated across the study period. The population of all provinces and territories, except Quebec, by age groups was used to calculate rates per 100,000 population. The 18-49 years age group was included in the analysis for risk factors of interest. Data on risk factors were aggregated across age groups and seasons. Descriptive data analyses were performed in SAS 9.4 and figures were produced using Microsoft Excel 365.The results from the analysis of Canadian hospitalization data were compared with the evidence from the rapid review. Results from both sources are presented and summarized by outcome of interest, except for medically attended RSV RTI for which data was only available from the rapid review.

## Results

After deduplication, 1,313 references were screened for study eligibility in the rapid review (Figure 1). Overall, 26 articles, including 7 SRs, were incorporated into the narrative synthesis of RSV burden of disease in adults (Table 1).

**Figure 1:**
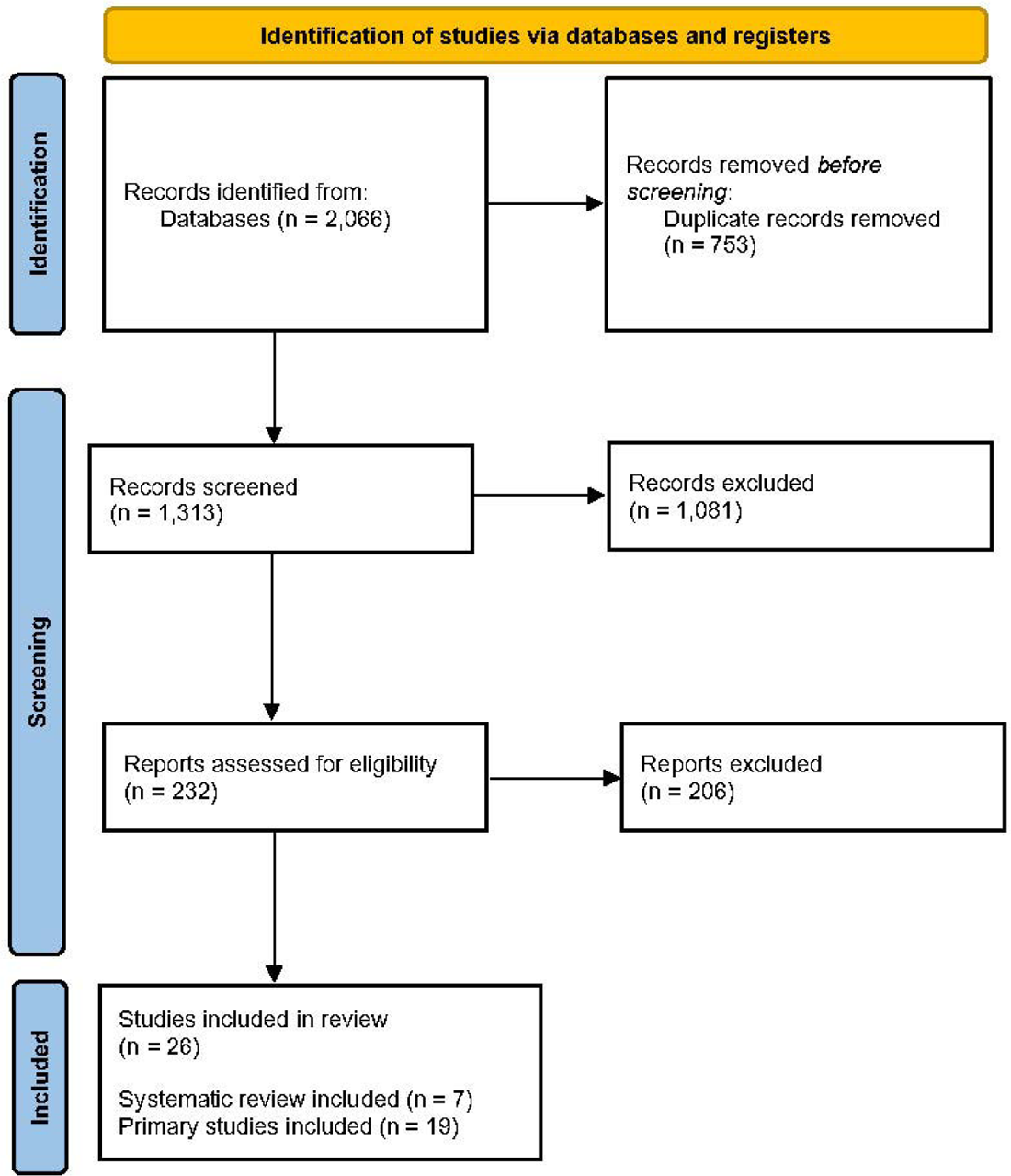
Study selection Prisma flow diagram

**Table 1:**
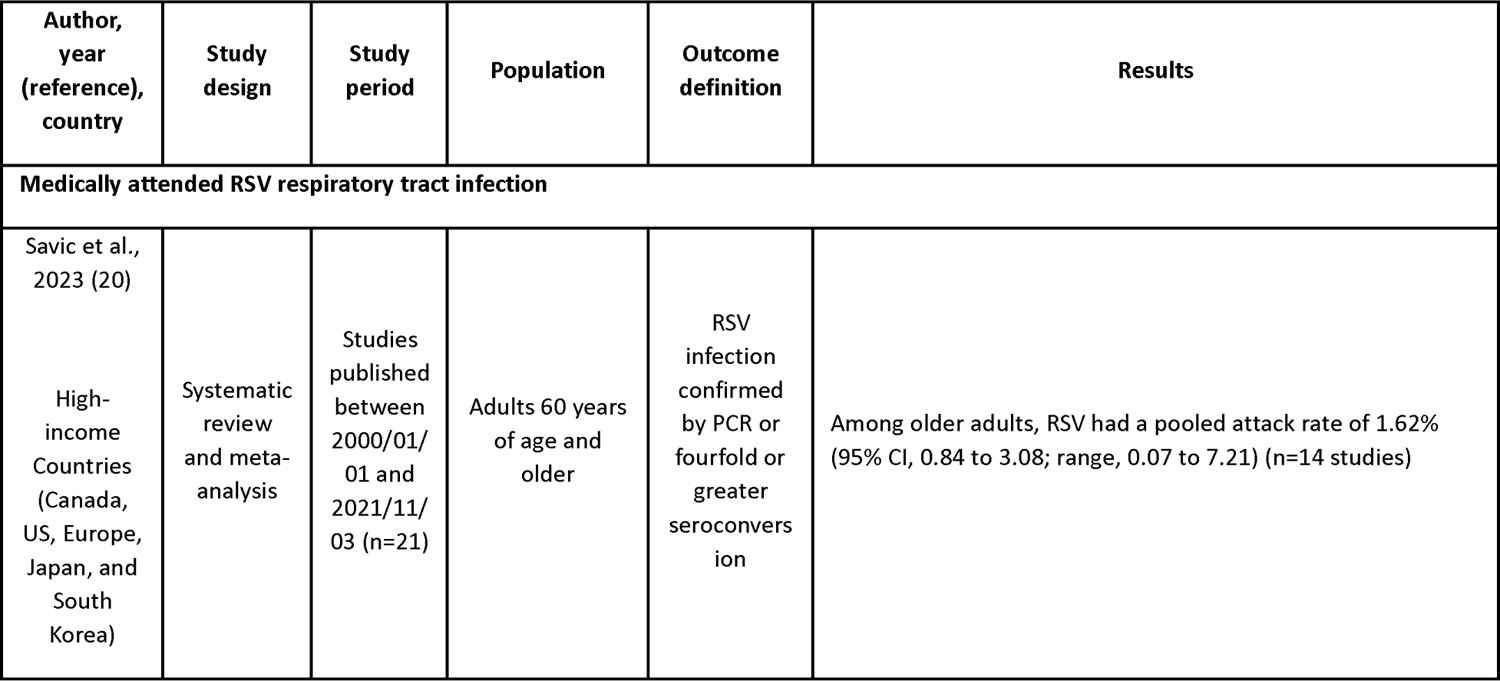
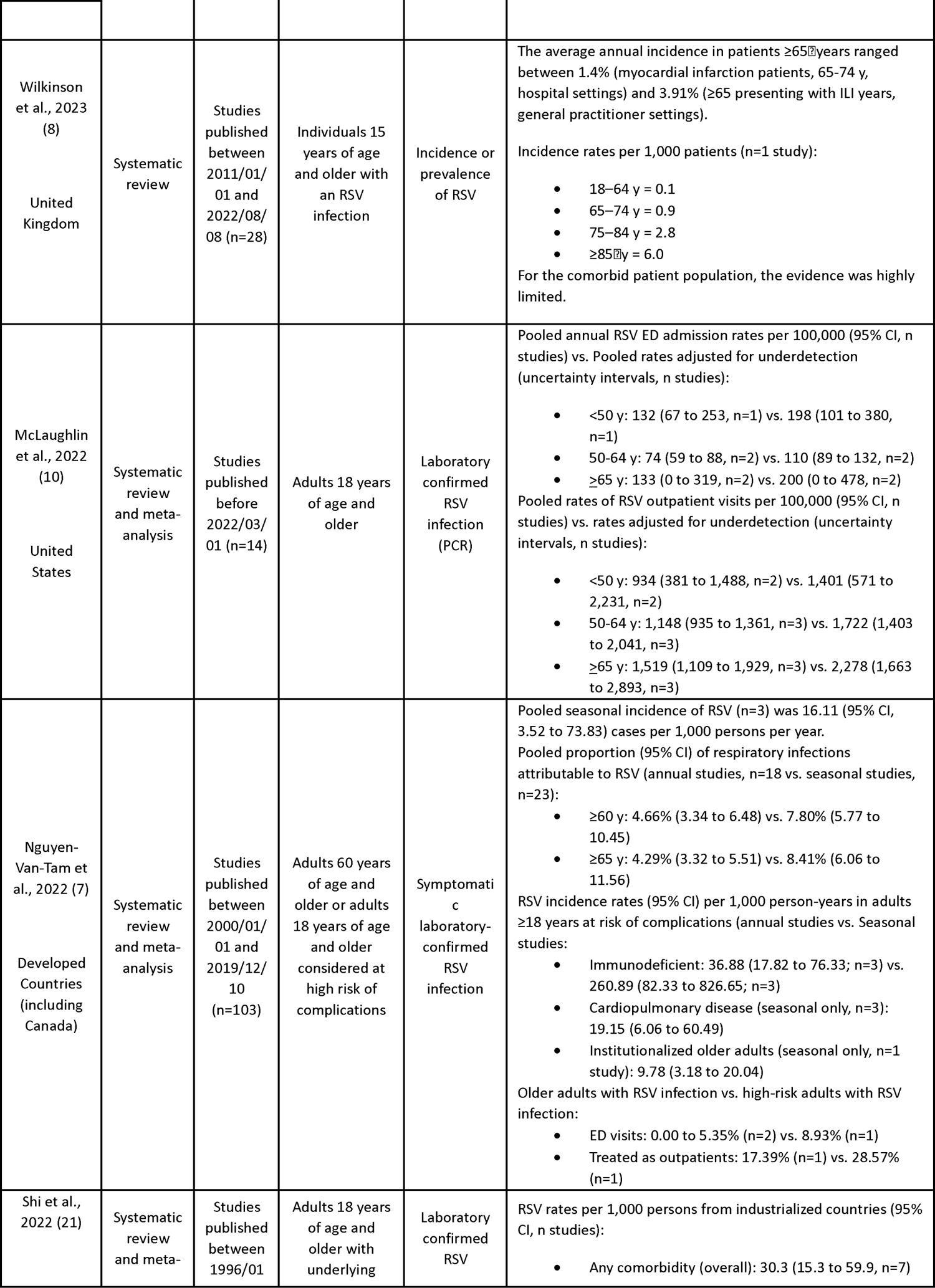
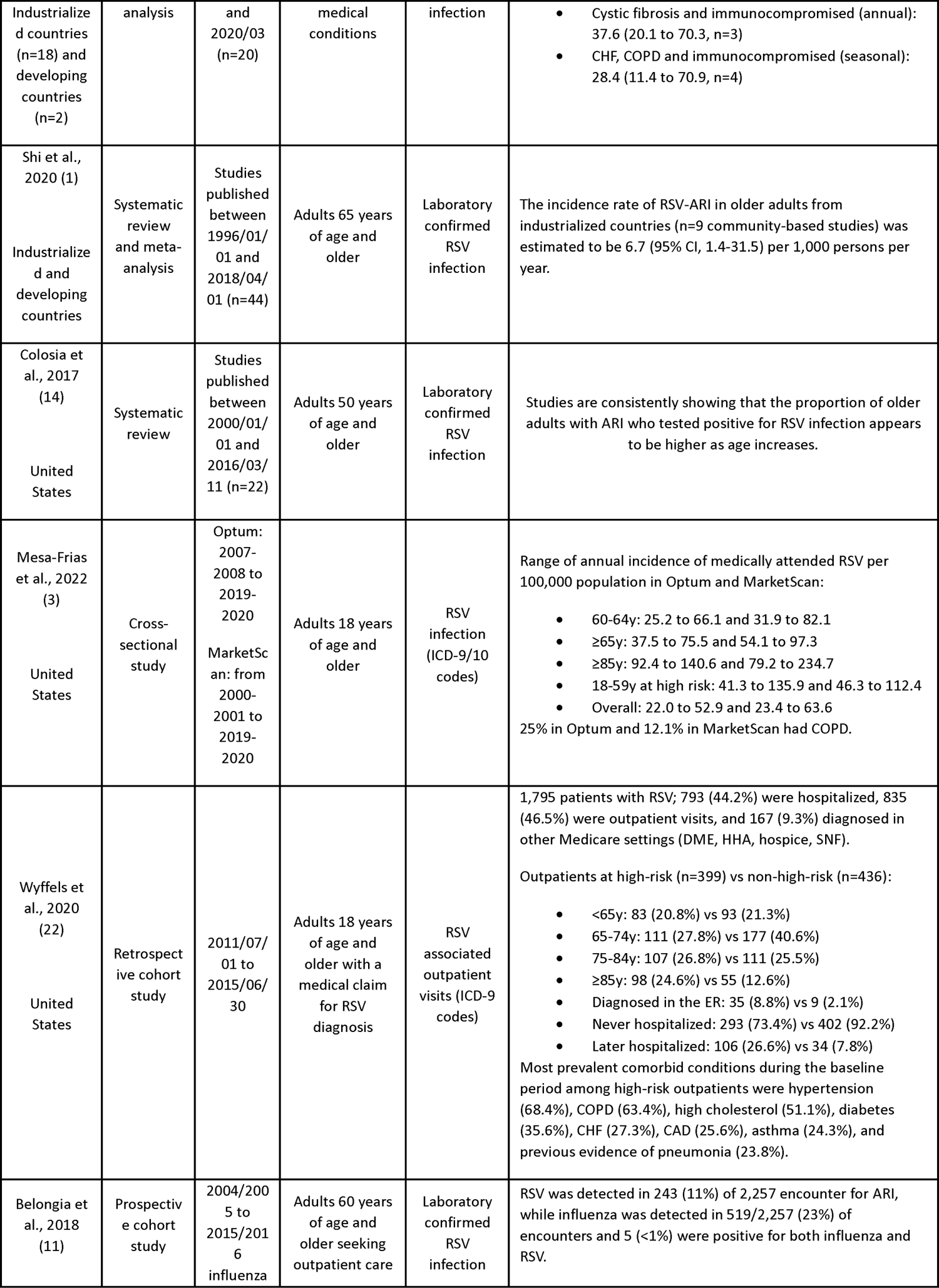
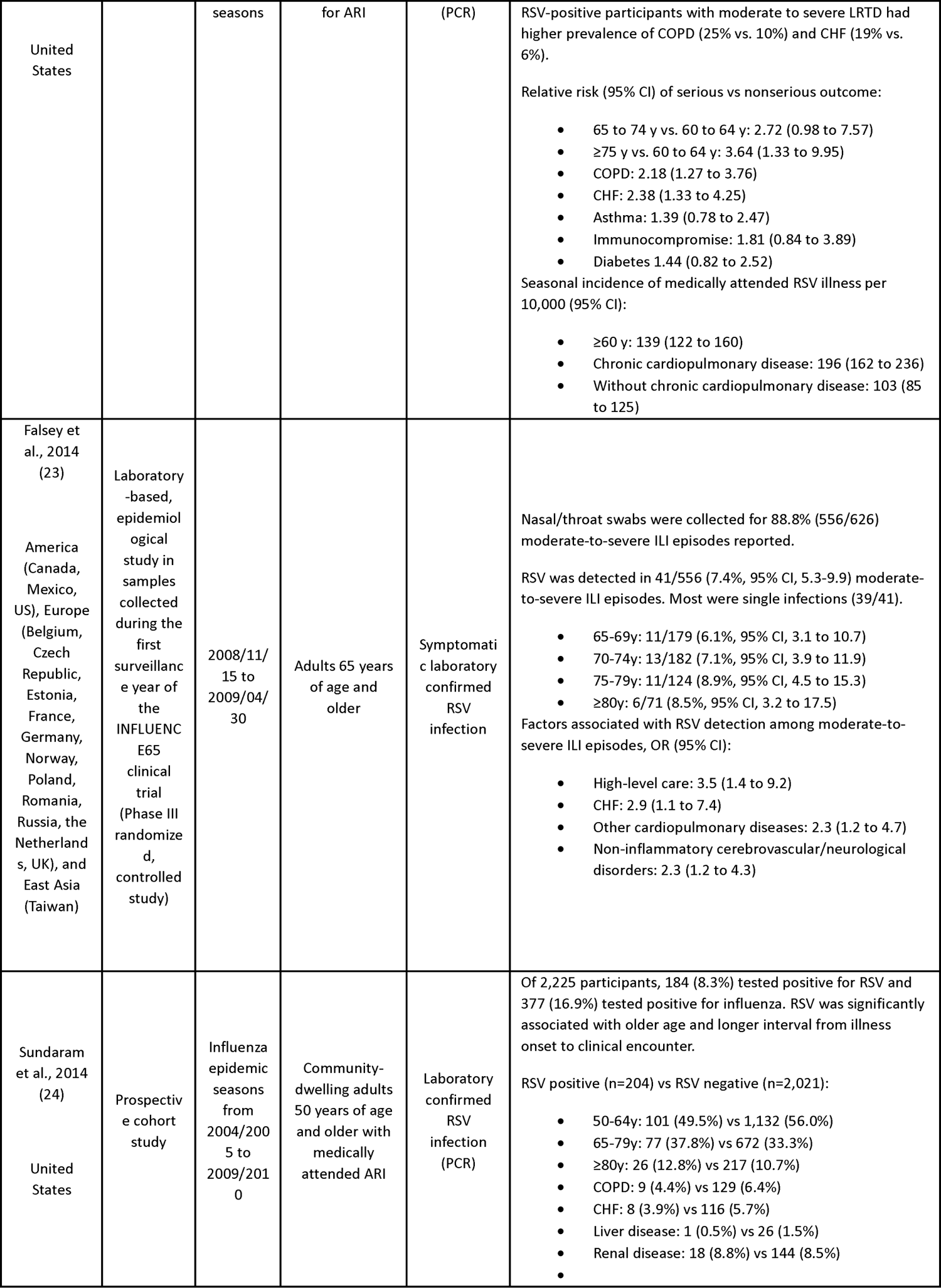
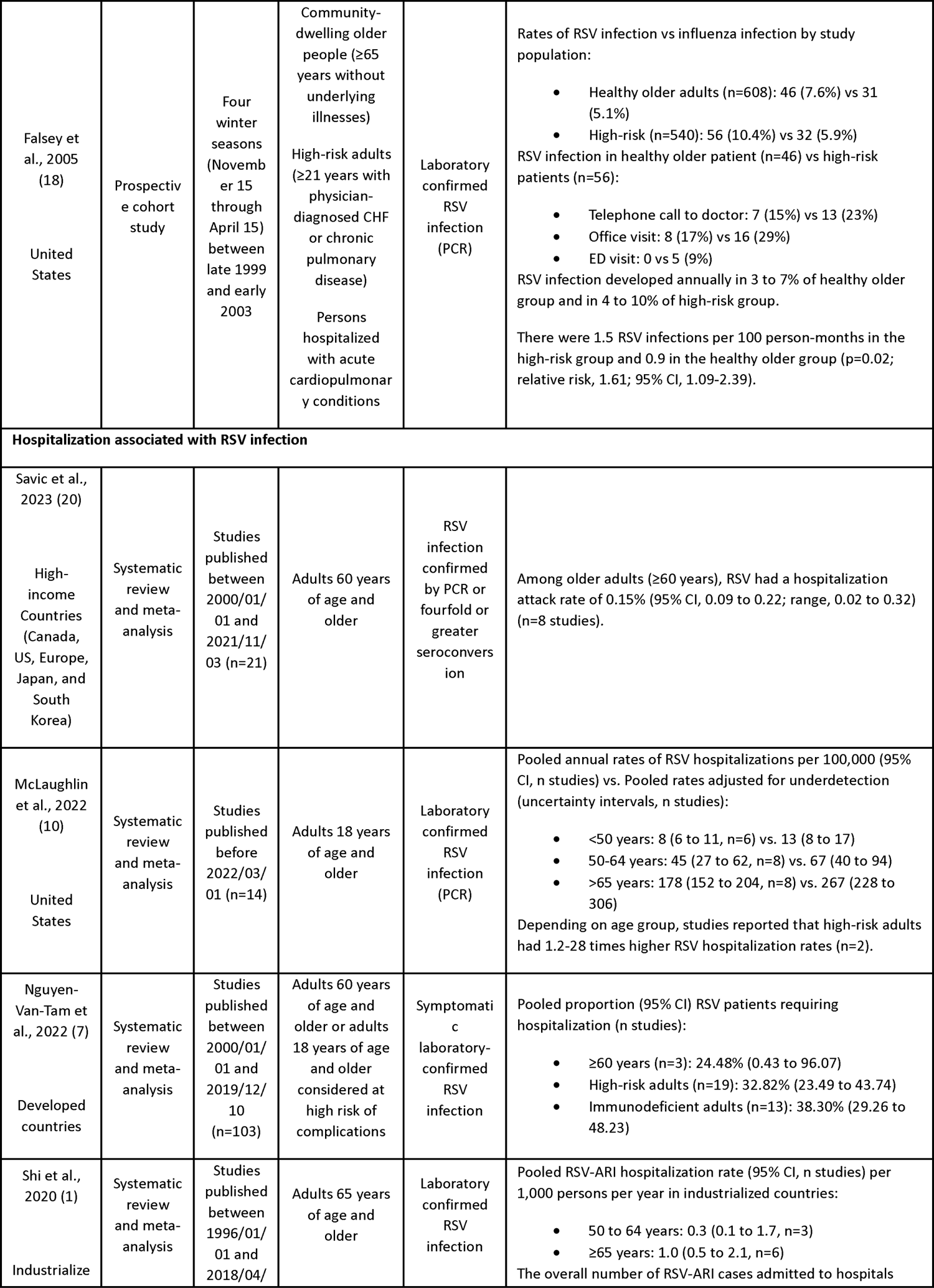
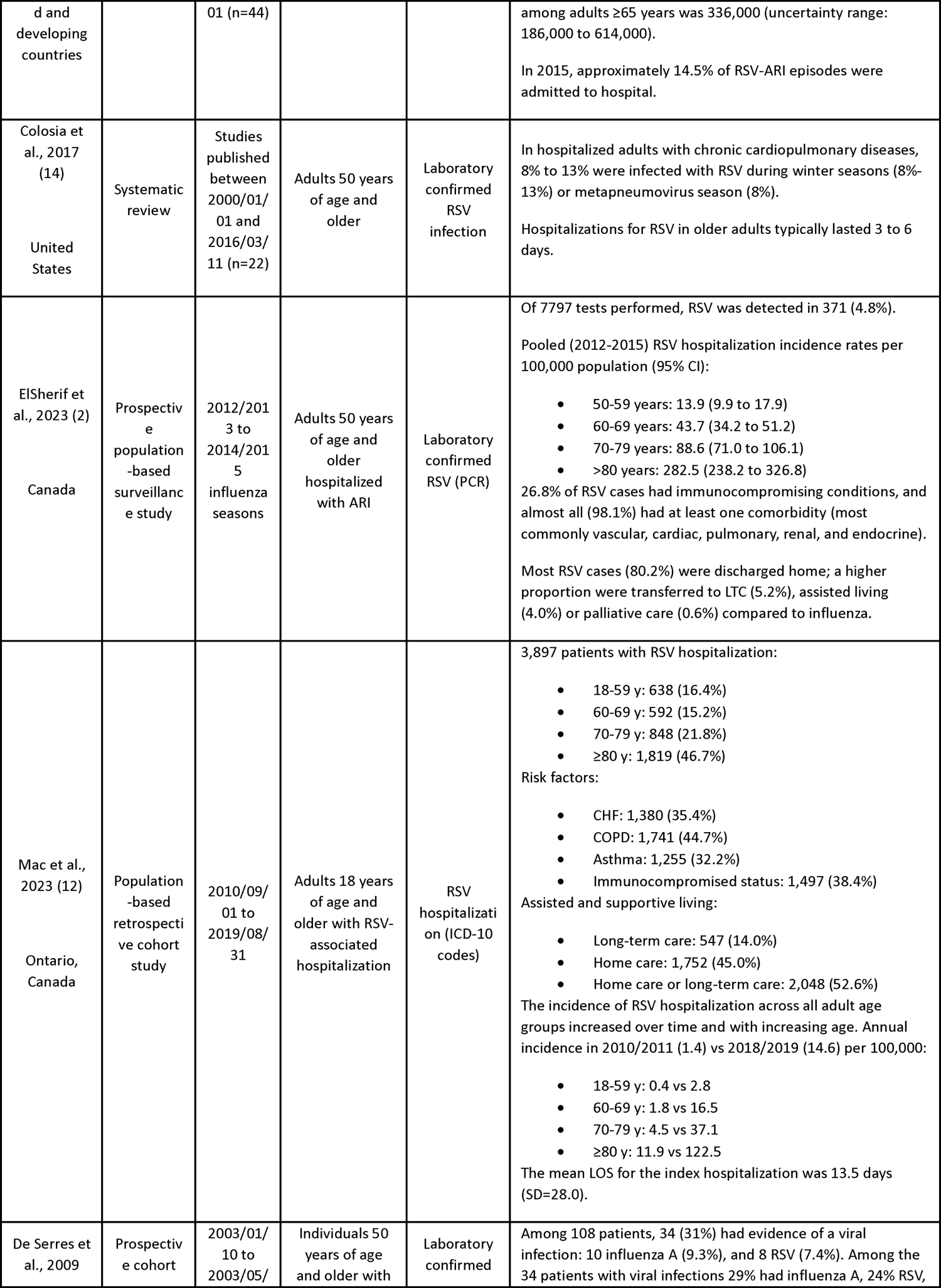
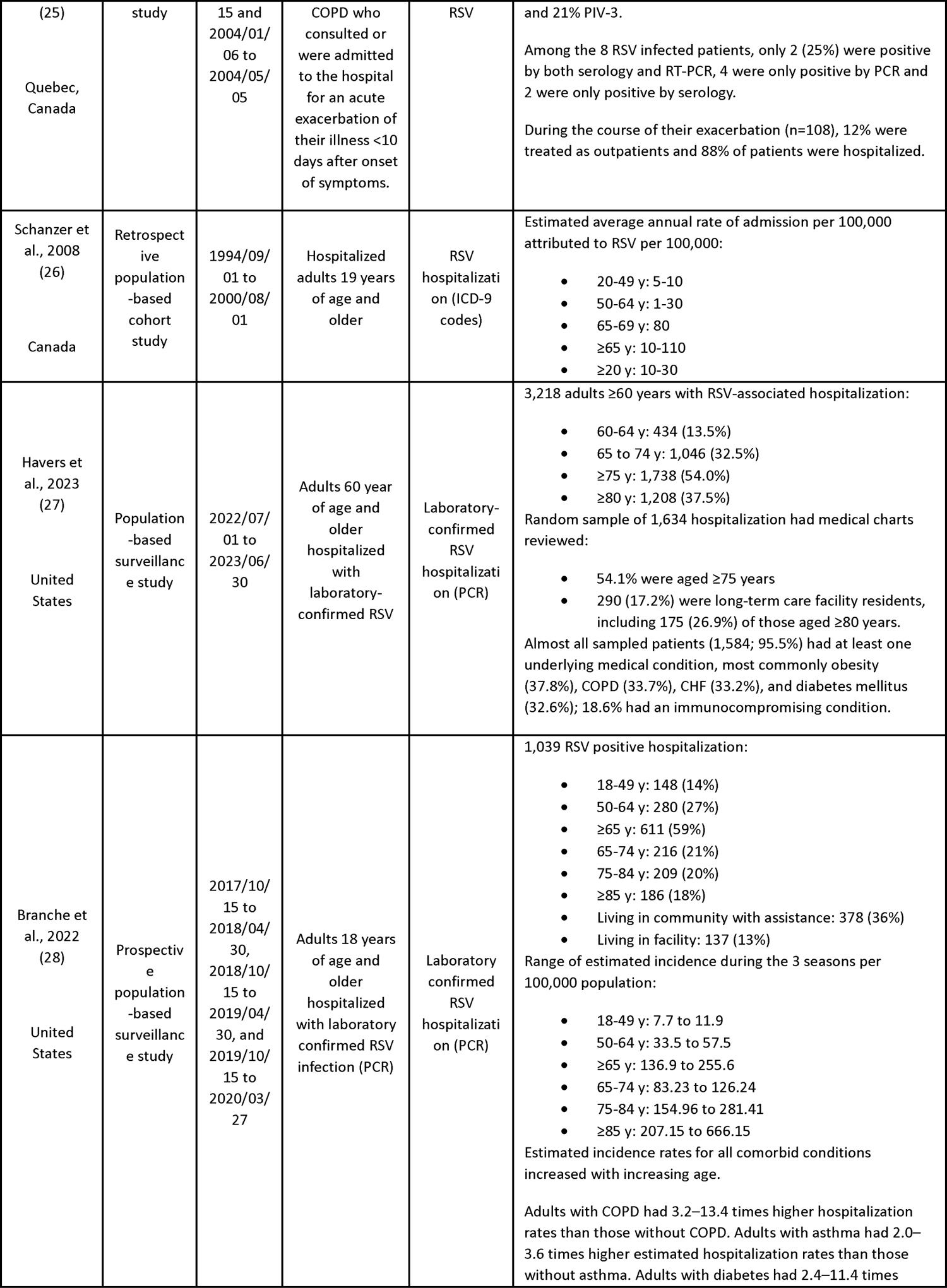
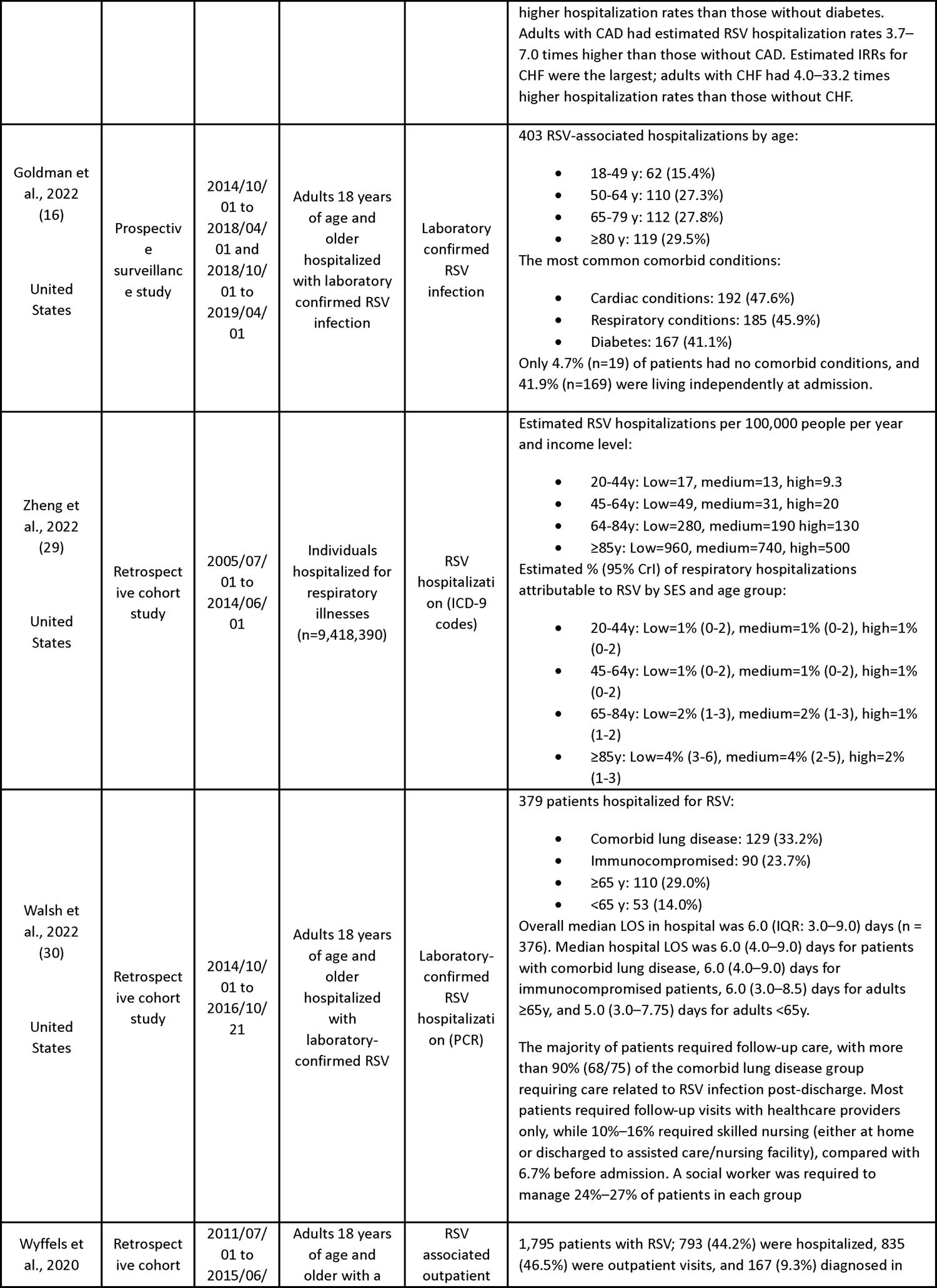
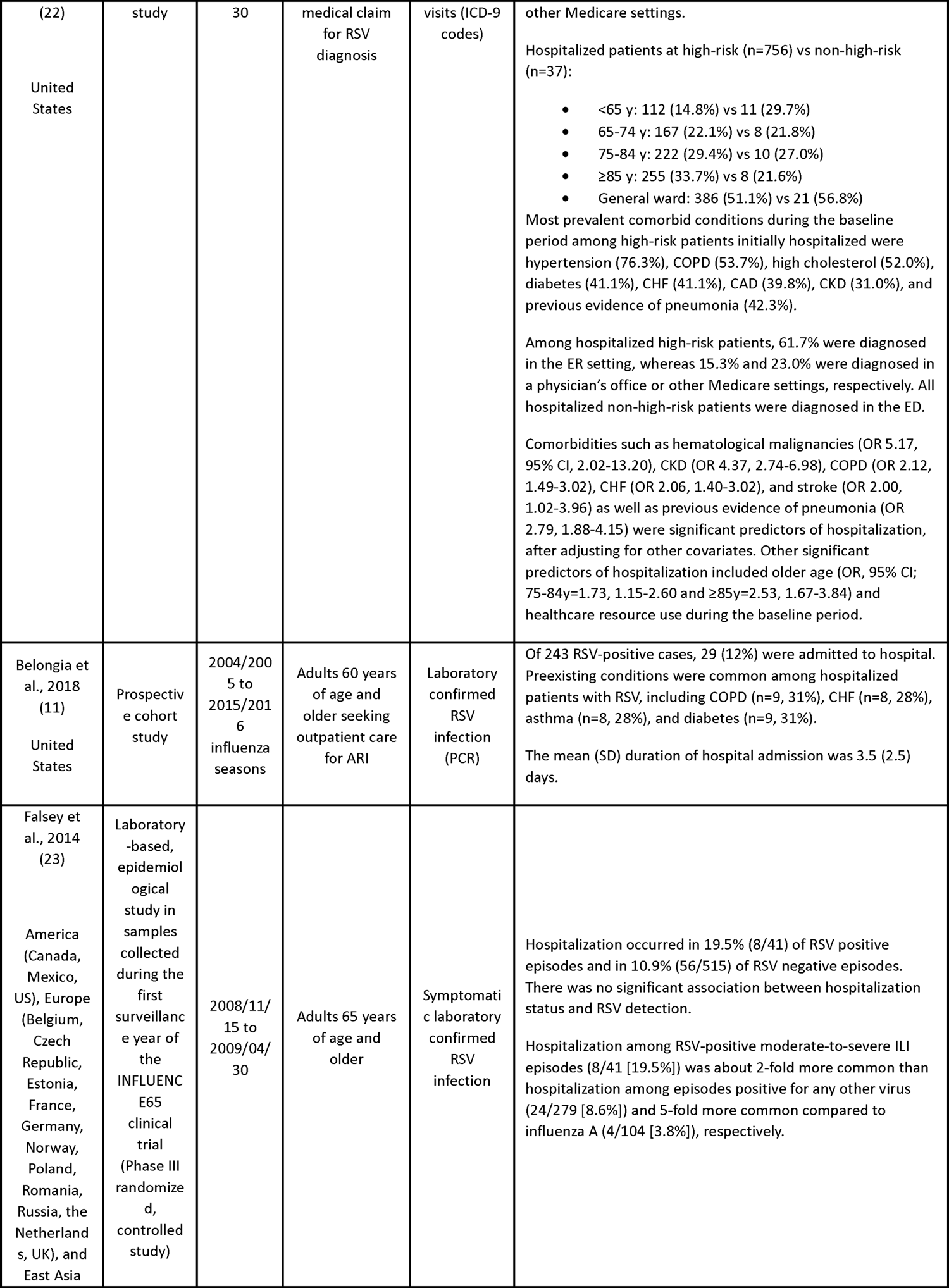
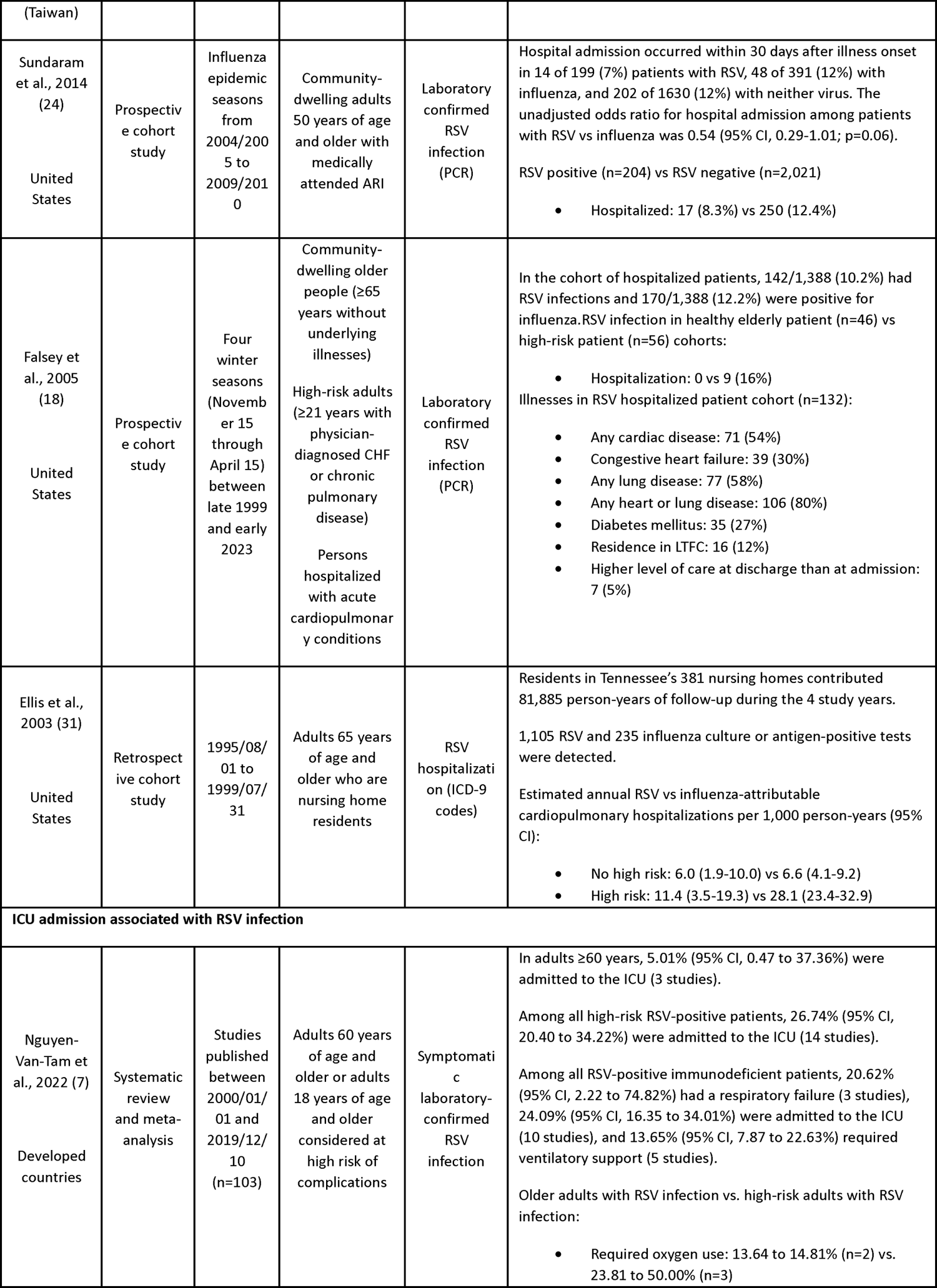
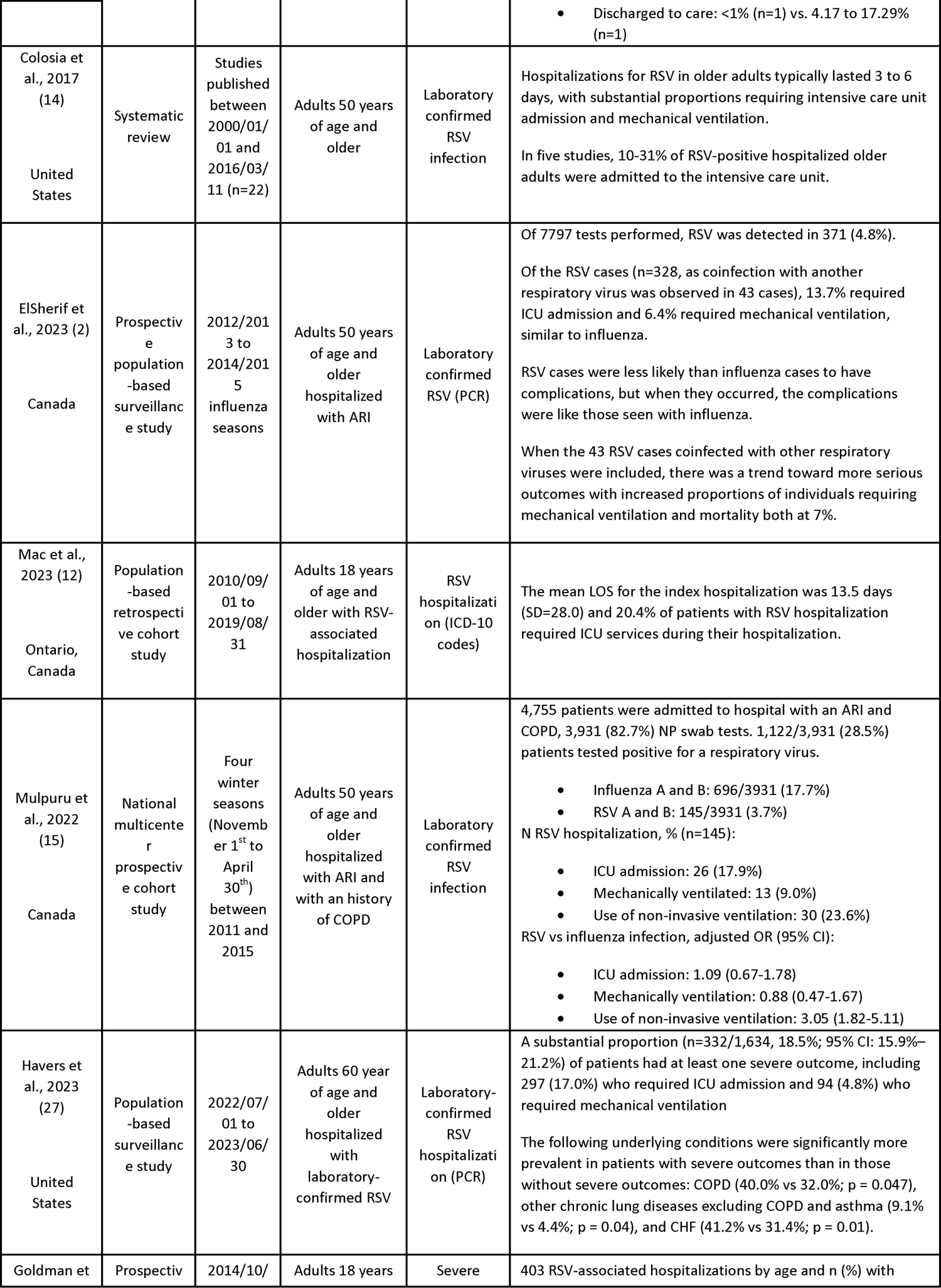
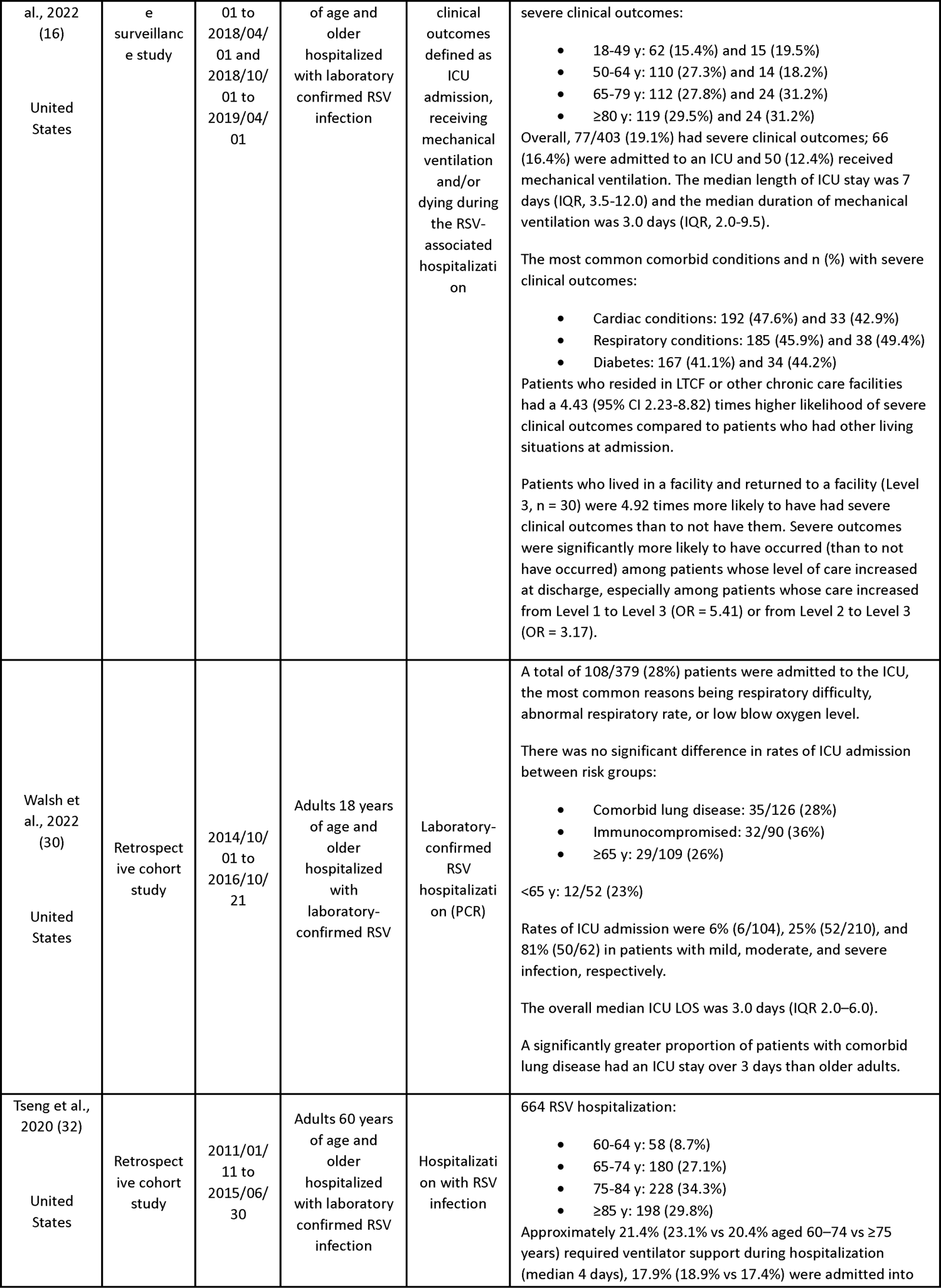
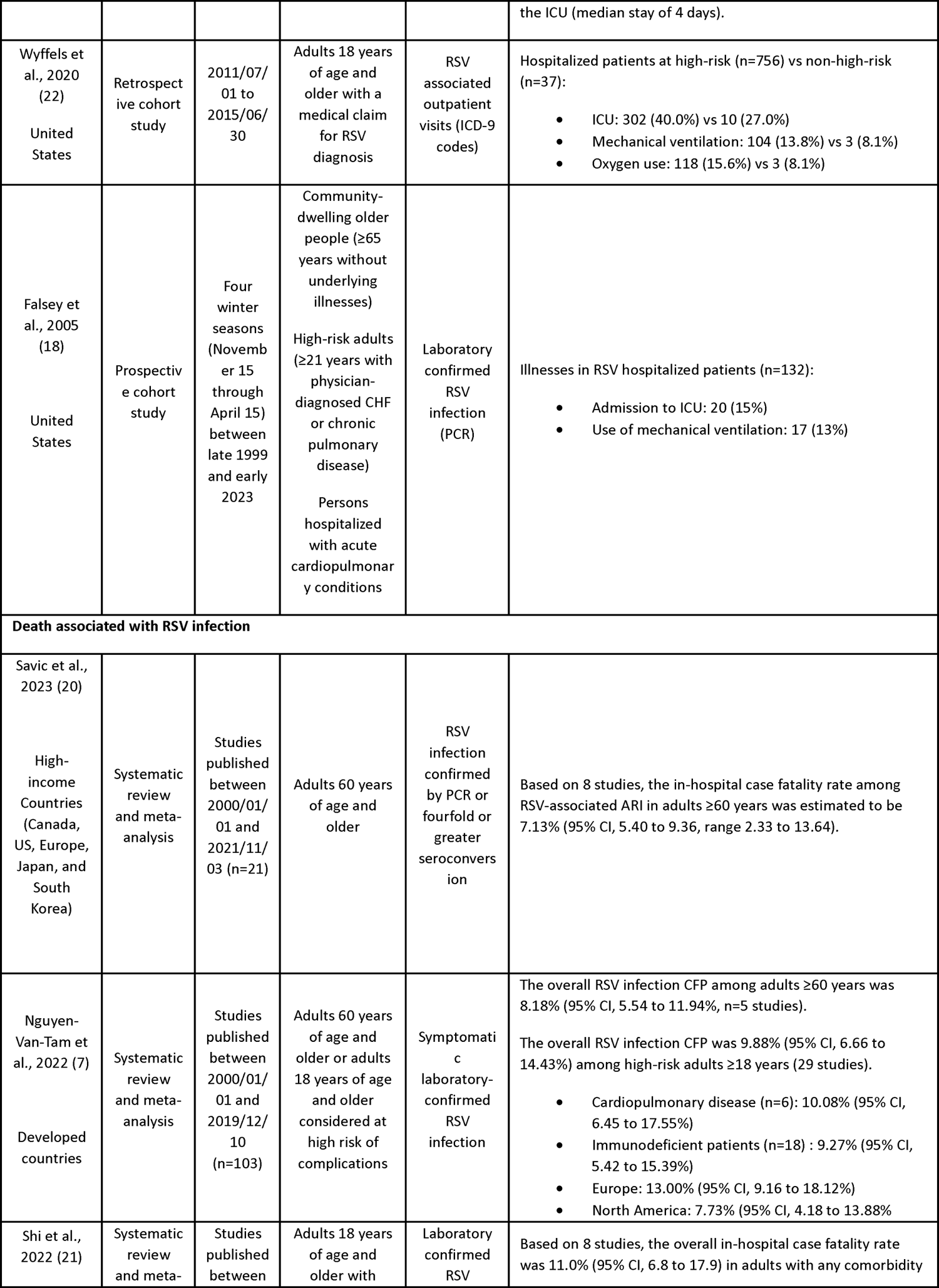
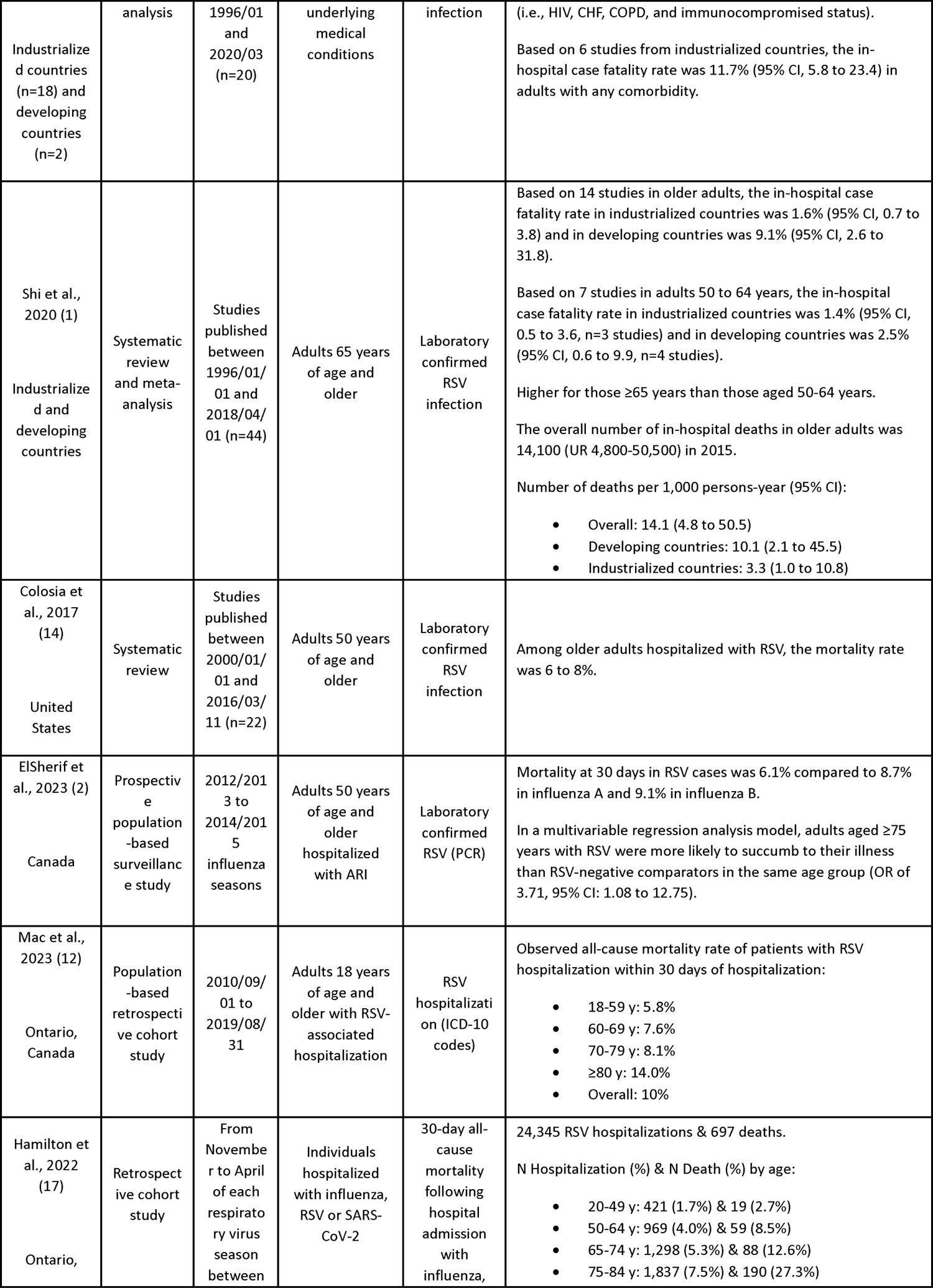
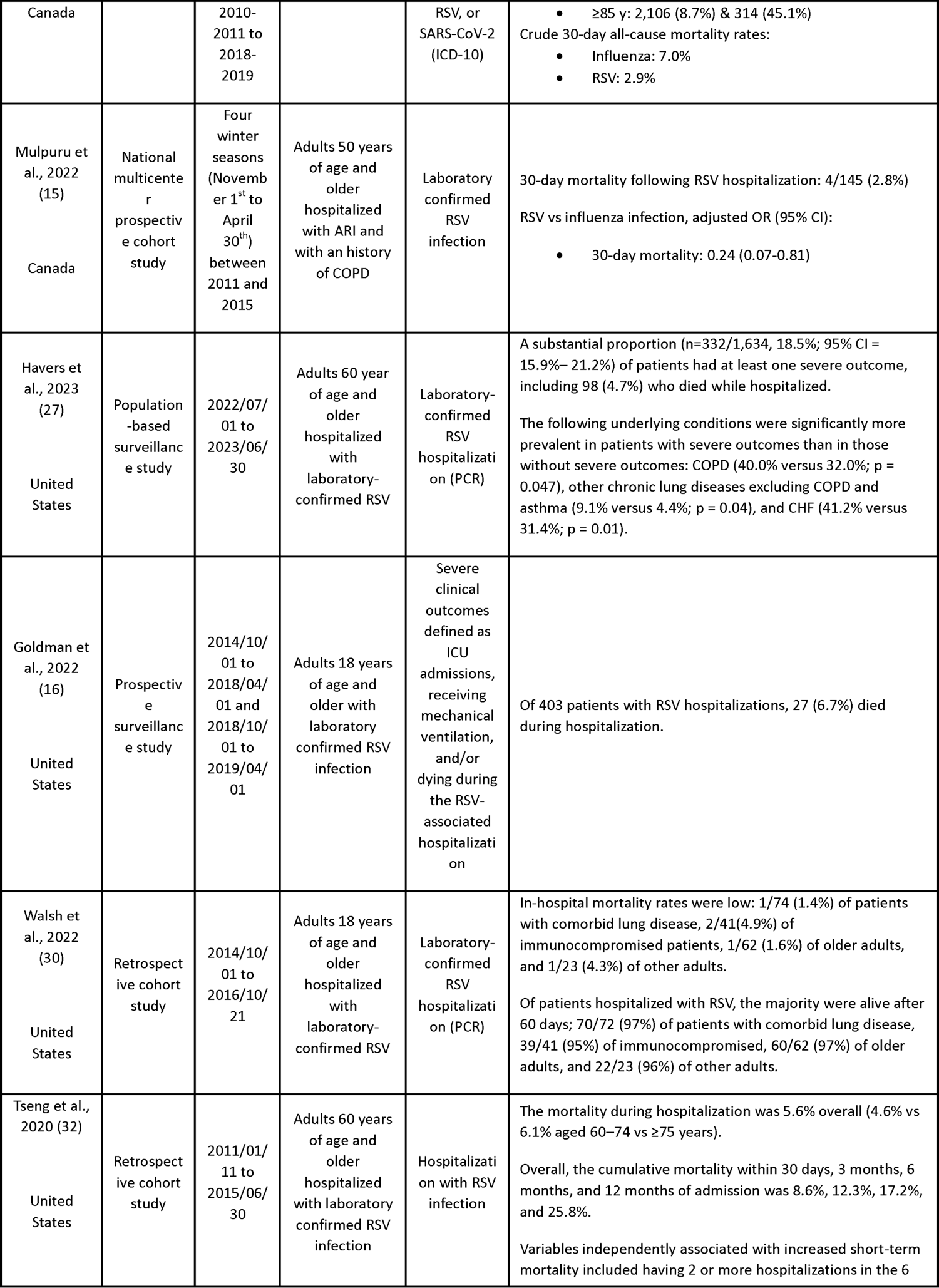
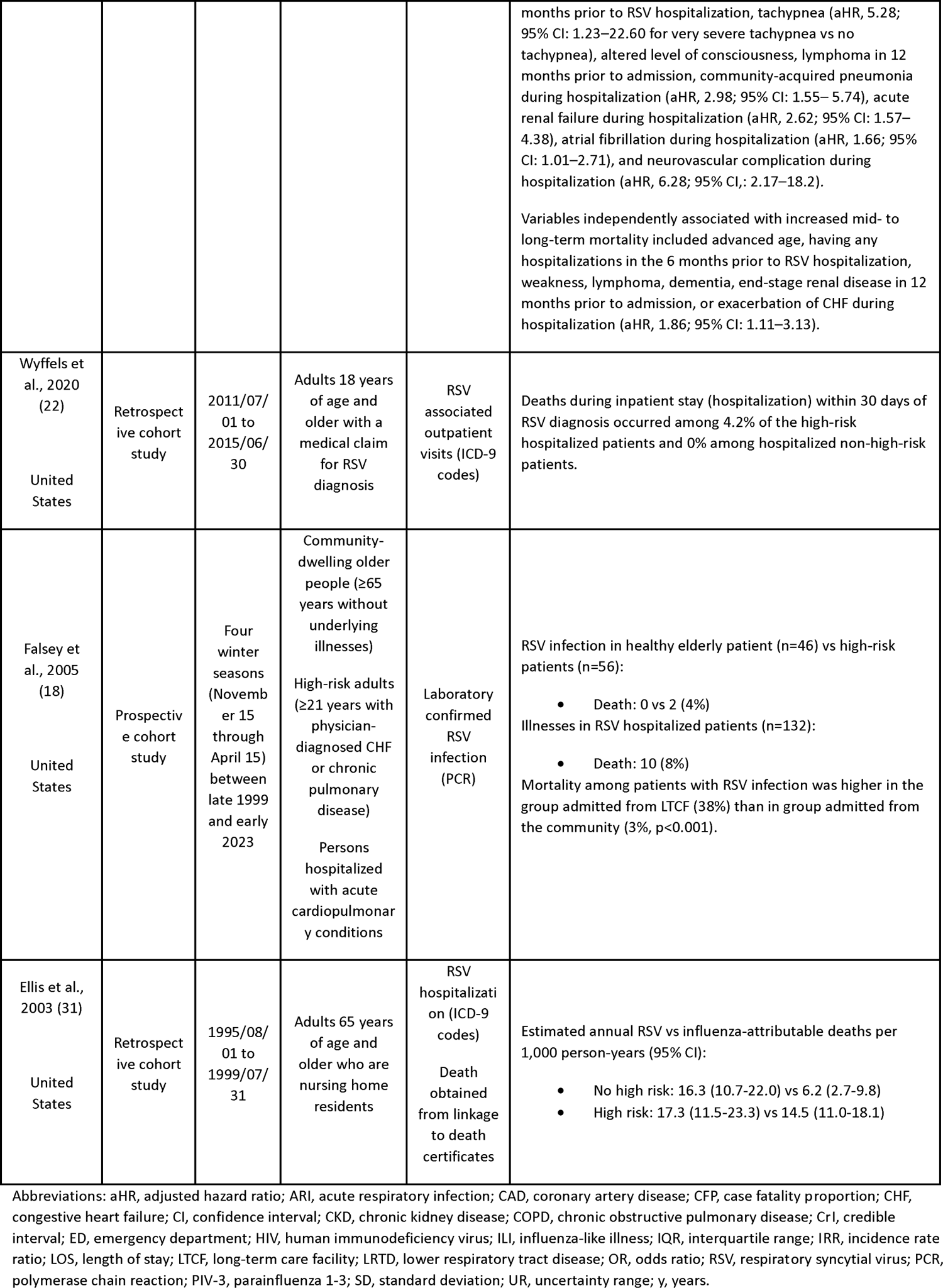
Summary of included studies on the burden of disease of respiratory syncytial virus in adults.

### Medically Attended RSV Respiratory Tract Infection

#### Rapid Review

Seven SRs and six observational studies describing the incidence of medically attended RSV RTI in older adults as well as adults with underlying medical conditions were identified; some included Canadian data (n=3), but none were restricted to Canada (Table 1). In adults 60 years of age and older, a SR of developed countries, including Canada, found that RSV caused between 4.7% and 7.8% of symptomatic respiratory infections (7). Overall, the incidence of medically attended RSV RTI increased with increasing age (8, 9). For instance, a SR and meta-analysis (MA) found that rates of medically attended RSV RTI among adults from the United States (US) increased from 934 per 100,000 in adults 18 to 49 years of age to 1,519 per 100,000 in adults 65 years of age and older (10). Factors associated with severe RSV infection in adults 65 years of age and older included increasing age and the presence of underlying medical conditions (i.e., cardiorespiratory disease, diabetes, and immunocompromising conditions). In a prospective US cohort study of adults 60 years of age and older, incidence was almost two times higher among adults with chronic cardiopulmonary disease compared to those without (incidence rate ratio (IRR) of 1.89, 95% confidence interval (CI): 1.44-2.48) (11). Although evidence was limited, studies suggest that RSV incidence is high in younger adults (i.e., 18 to 59 years) with certain medical conditions and is somewhat similar to adults 65 years of age and older. A cross-sectional study from the US of the annual incidence of medically attended RSV found that incidence was highest in adults 85 years of age and older, followed by 65 years of age and older, and then followed closely by 18 to 59 years of age considered at high risk of severe RSV including those with cardiorespiratory disease or immunocompromising conditions (3).

### Hospitalization Associated with RSV Infection

#### Rapid Review

Six SRs and fifteen observational studies, including four Canadian studies, described the incidence of hospitalization associated with RSV infection. In general, studies found that the incidence increased consistently with increasing age. For instance, a prospective Canadian population-based surveillance study found the following average seasonal RSV hospitalizations incidence rates per 100,000 between 2012-2015: 13.9 (95% CI: 9.9-17.9) in adults 50 to 59 years, 43.7 (95% CI: 34.2-51.2) in those 60 to 69 years, 88.6 (95% CI: 71.0-106.1) in adults 70 to 79 years, and 282.5 (95% CI: 238.2-326.8) in adults 80 years of age and older (2). A SR found that depending on age and risk factors, adults 18 years of age and older with chronic medical conditions have higher rates of hospitalization associated with RSV compared to those without the condition (10). The authors reported rates ranging from 1.2 to 1.3 times higher for adults with obesity to 27.6 times higher for those 20 to 39 years of age with congestive heart failure (CHF) (10). Similarly, a retrospective cohort study from Ontario found that among adults 18 years of age and older who had a hospitalization associated with RSV between September 2010 and August 2017, 35.4% had CHF, 44.7% had COPD, 32.2% had asthma, and 38.4% had immunocompromising conditions (12). Another Canadian study found that 26.8% of adults 50 years of age and older who had a hospitalization associated with RSV over the 2012-2015 seasons had immunocompromising conditions and almost all (98.1%) had at least one comorbidity with the most frequent being vascular (71.3%), cardiac (55.5%), pulmonary (48.2%), renal (48.2%) and endocrine (33.2%) conditions (2).

#### Canadian Hospitalization Data

Across 12 seasons, there were a total of 19,436 recorded hospitalizations associated with RSV among adults 50 years of age and older, of which 6,314 were due to RSV, corresponding to an average of 1,630 hospitalizations associated with RSV per season (Table 2). Rates of hospitalizations associated with RSV among older adults in Canada generally increased over time across all age groups (Figure 2A). Overall, the average rate of hospitalization associated with RSV among adults 50 years of age and older was 16 per 100,000 population and the average rate of hospitalization associated with RSV per 100,000 population by age groups were the following: 4 in adults 50 to 59 years old, 10 in adults 60 to 69 years old, 22 in 70 to 79 years, and 63 in adults 80 years of age and older. The rates of hospitalizations due to RSV among older adults in Canada followed the same trend; however, rates were much lower (Figure 2B). In general, both rates of hospitalization associated with and due to RSV increased with increasing age (6, 13).

**Figure 2.**
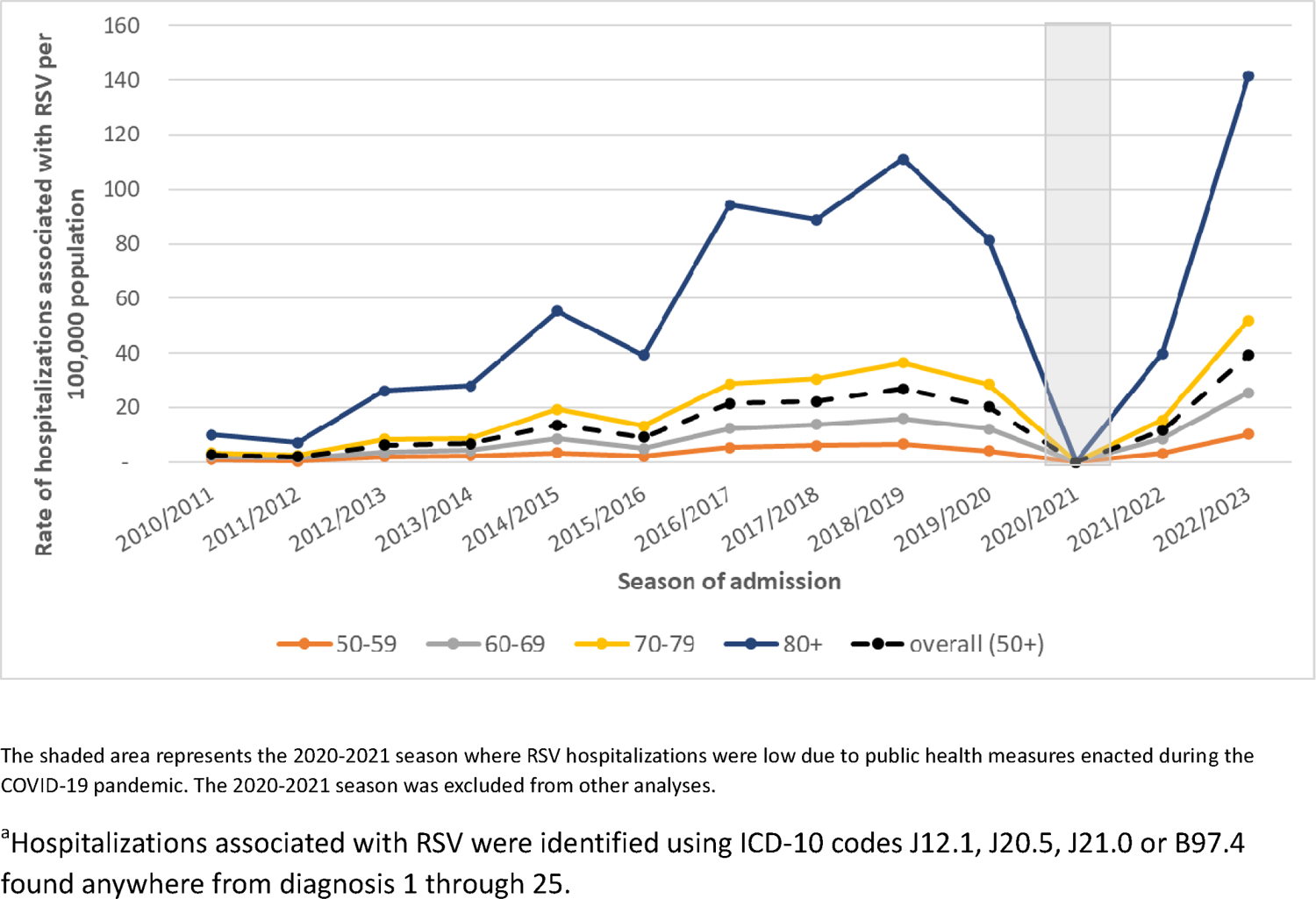
A) Rate of hospitalizations associated with ^a^ RSV, by age group (years), seasons 2010-2011 to 2019-2020 and 2021-2022 to 2022-2023, Canada (excluding Quebec), Canadian Discharge Abstract Database

**Figure 2.**
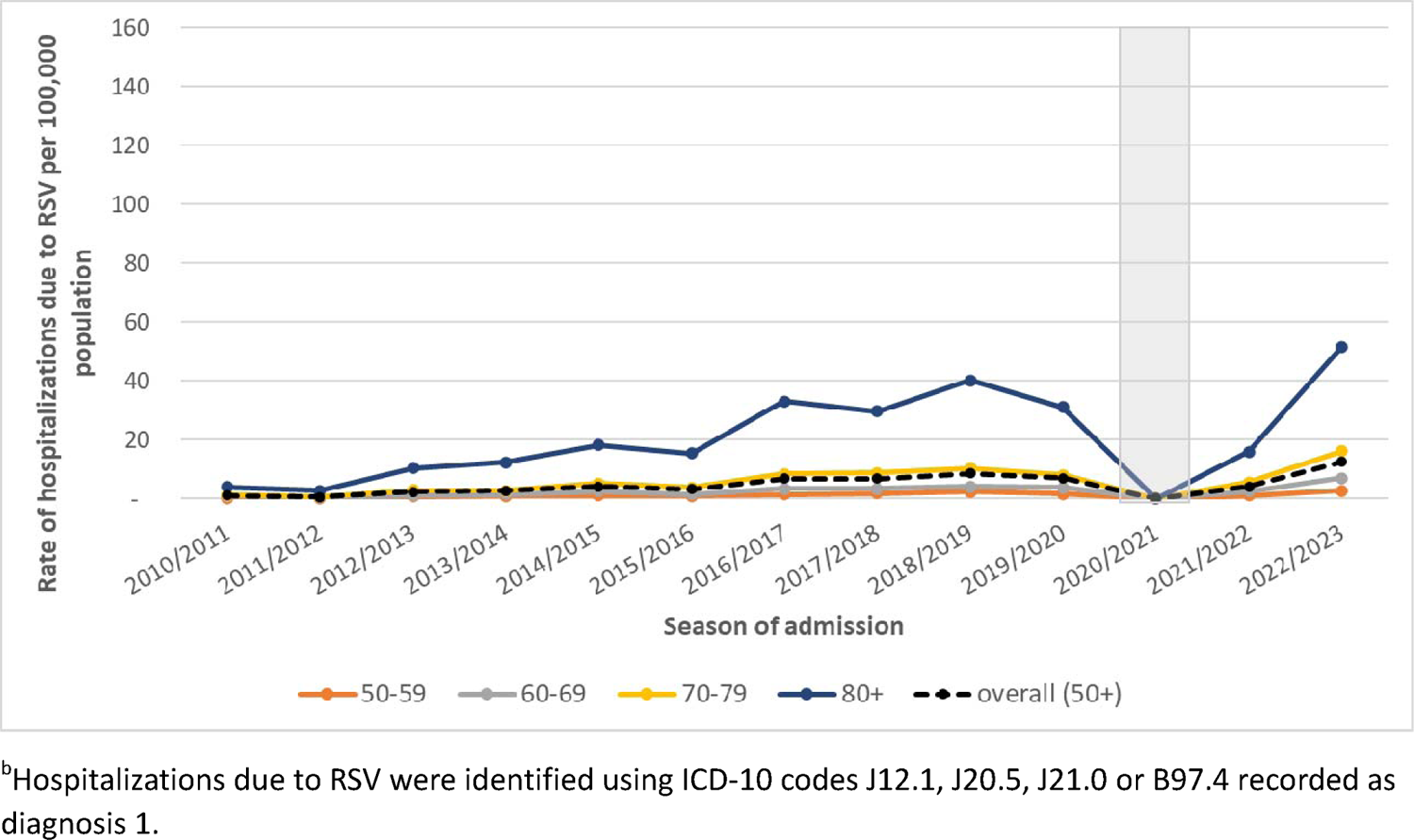
B) Rate of hospitalizations due to ^b^ RSV, by age group (years), seasons 2010-2011 to 2019-2020 and 2021-2022 to 2022-2023, Canada (excluding Quebec), Canadian Discharge Abstract Database

**Table 2:**
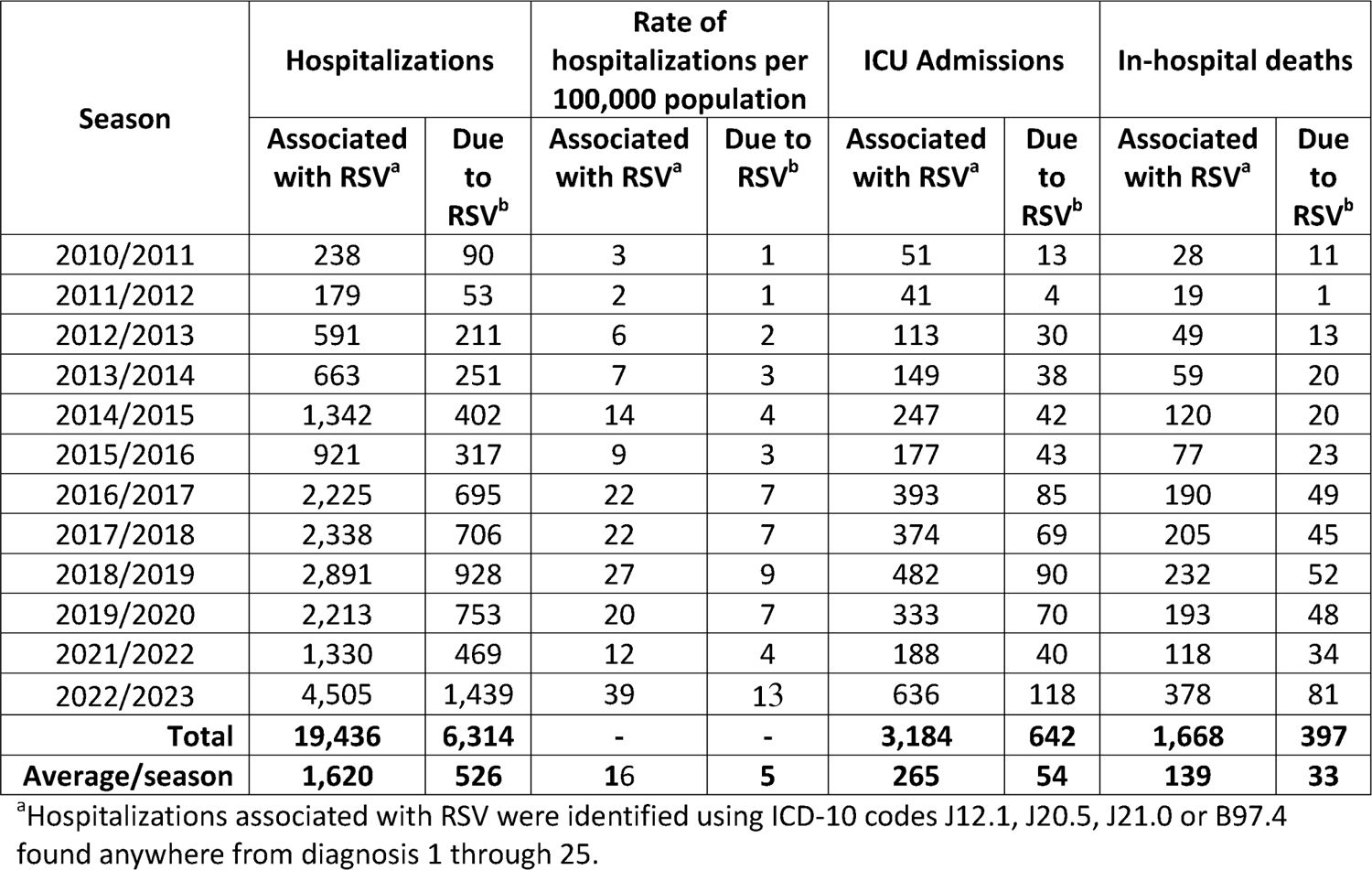
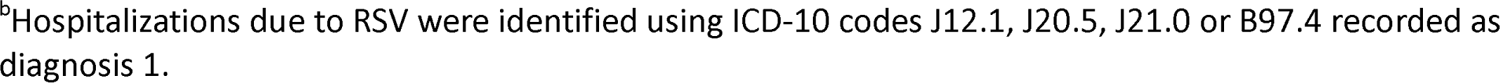
Total hospitalizations, ICU admissions and in-hospital deaths associated with and due to RSV, adults aged 50 years of age and older, seasons 2010-2011 to 2019-2020 and 2021-2022 to 2022-2023, Canada (excluding Quebec), Canadian Discharge Abstract Database.

| Season | Hospitalizations |  | Rate of hospitalizations per 100,000 population |  | ICU Admissions |  | In-hospital deaths |  |
| --- | --- | --- | --- | --- | --- | --- | --- | --- |
|  | Associated with RSV<sup>a</sup> | Due to RSV<sup>b</sup> | Associated with RSV<sup>a</sup> | Due to RSV<sup>b</sup> | Associated with RSV<sup>a</sup> | Due to RSV<sup>b</sup> | Associated with RSV<sup>a</sup> | Due to RSV<sup>b</sup> |
| 2010/2011 | 238 | 90 | 3 | 1 | 51 | 13 | 28 | 11 |
| 2011/2012 | 179 | 53 | 2 | 1 | 41 | 4 | 19 | 1 |
| 2012/2013 | 591 | 211 | 6 | 2 | 113 | 30 | 49 | 13 |
| 2013/2014 | 663 | 251 | 7 | 3 | 149 | 38 | 59 | 20 |
| 2014/2015 | 1,342 | 402 | 14 | 4 | 247 | 42 | 120 | 20 |
| 2015/2016 | 921 | 317 | 9 | 3 | 177 | 43 | 77 | 23 |
| 2016/2017 | 2,225 | 695 | 22 | 7 | 393 | 85 | 190 | 49 |
| 2017/2018 | 2,338 | 706 | 22 | 7 | 374 | 69 | 205 | 45 |
| 2018/2019 | 2,891 | 928 | 27 | 9 | 482 | 90 | 232 | 52 |
| 2019/2020 | 2,213 | 753 | 20 | 7 | 333 | 70 | 193 | 48 |
| 2021/2022 | 1,330 | 469 | 12 | 4 | 188 | 40 | 118 | 34 |
| 2022/2023 | 4,505 | 1,439 | 39 | 13 | 636 | 118 | 378 | 81 |
| **Total** | **19,436** | **6,314** | - | - | **3,184** | **642** | **1,668** | **397** |
| **Average/season** | **1,620** | **526** | **16** | **5** | **265** | **54** | **139** | **33** |

<sup>a</sup>Hospitalizations associated with RSV were identified using ICD-10 codes J12.1, J20.5, J21.0 or B97.4 found anywhere from diagnosis 1 through 25.
<sup>b</sup>Hospitalizations due to RSV were identified using ICD-10 codes J12.1, J20.5, J21.0 or B97.4 recorded as diagnosis 1.

A total of 21,258 hospitalizations associated with RSV among adults 18 years of age and older were reported across the 12 seasons (Table 3). Of these hospitalizations, 76.4% were reported to have at least one risk factor of interest, 34.6% were reported to have at least two of these risk factors and 9.1% were reported to have at least three of these risk factors. Among these 21,258 hospitalizations, 30.0% reported having COPD, 29.6% had diabetes, 23.4% had cardiovascular disease, 16.8% had an immunocompromising condition, 15.8% had a respiratory tract infection and 6.3% had chronic kidney disease.

**Table 3:** Number and percent hospitalizations associated with RSV with risk factor of interest, adults aged 18 years of age and older, seasons 2010-2011 to 2019-2020 and 2021-2022 to 2022-2023, Canada (excluding Quebec), Canadian Discharge Abstract Database.

| **Risk Factor of Interest** | **Number of Hospitalizations<sup>a</sup>** | **Percentage (%)<sup>b</sup>** |
| --- | --- | --- |
| Chronic obstructive pulmonary disease | 6,360 | 30.0 |
| Diabetes | 6,276 | 29.6 |
| Cardiovascular Disease | 4,965 | 23.4 |
| Immunosuppressive Conditions | 3,564 | 16.8 |
| Respiratory tract infection | 3,344 | 15.8 |
| Chronic Kidney Disease | 1,336 | 6.3 |
| Total number of hospitalizations associated with RSV<sup>c</sup> | 21,258 |  |
| **Total Number of risk factors of interest** |  |  |
| At least 1 | 16,250 | 76.4 |
| 2 or more | 7,345 | 34.6 |
| 3 or more | 1,928 | 9.1 |
| 4 or more | 297 | 1.4 |

<sup>a</sup>Number of hospitalizations by risk factor will not equal 21,258 hospitalizations as one patient may have multiple risk factors and the occurrence of each risk factor was considered mutually exclusive
<sup>b</sup>Percentage of hospitalizations will exceed 100% as one patient may have multiple risk factors and the occurrence of each risk factor was considered mutually exclusive
<sup>c</sup>Hospitalizations associated with RSV were identified using ICD-10 codes J12.1, J20.5, J21.0 or B97.4 found anywhere from diagnosis 1 through 25.

#### ICU Admission Associated with RSV Infection Rapid Review

Two SRs and nine observational studies, including three Canadian studies, reported data on ICU admission associated with RSV infection. A Canadian prospective population-based surveillance study found that among adults 50 years of age and older hospitalized with RSV, 13.7% required ICU admission and 6.4% required mechanical ventilation (similar to influenza) between 2012 and 2015 (2). As with other clinical outcomes, risk increased with increasing age and the presence of comorbidities although granularity by age group is limited (7, 14). A SR from developed countries found a higher proportion of adults 18 years of age and older considered at risk of complications of infection was admitted to the ICU (26.7% versus 5.0%), required oxygen use (23.8-50.0% versus 13.6-14.8%), and was discharged to care (4.2-17.3% versus <1%) compared to adults 60 years of age and older (7). A Canadian prospective cohort study found that among adults 50 years of age and older with a history of COPD hospitalized with RSV during the winter seasons of 2011 to 2015, 17.9% required ICU admission, 9.0% were mechanically ventilated, and 23.6% needed non-invasive ventilation (15). A surveillance study from the US found that patients who resided in LTC or other chronic care facilities had a 4.43 (95% CI: 2.23-8.82) times higher likelihood of severe clinical outcomes (i.e., ICU admission, receiving mechanical ventilation and/or death) compared to patients with other living situations at admission (16).

#### Canadian Hospitalization Data

Across 12 seasons, among the 19,436 hospitalizations associated with RSV, 3,184 (16%) required ICU admission and among the 6,314 hospitalizations due to RSV, 642 (10%) required ICU admission. The average rate of ICU admissions associated with RSV among adults 50 years of age and older across the 12 seasons were 2.6 per 100,000 population and increased with increasing age (1.1 in adults 50 to 59 years, 2.4 in 60 to 69 years, 4.3 in 70 to 79 years, and 6.0 in adults 80 years of age and older) (Table 4). Rates of ICU admissions due to RSV in Canadian older adults followed the same trend; however, rates were much lower.

**Table 4:**
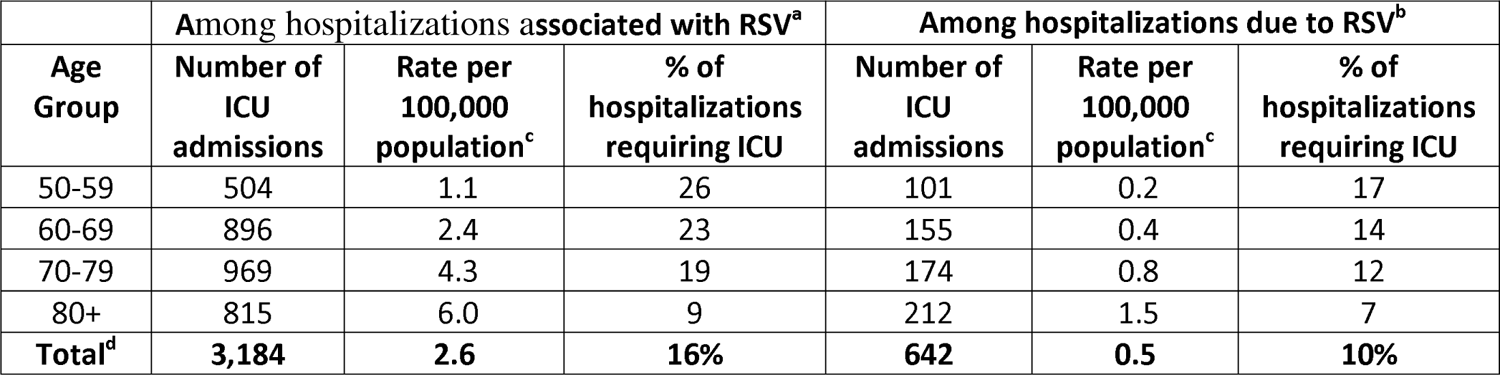
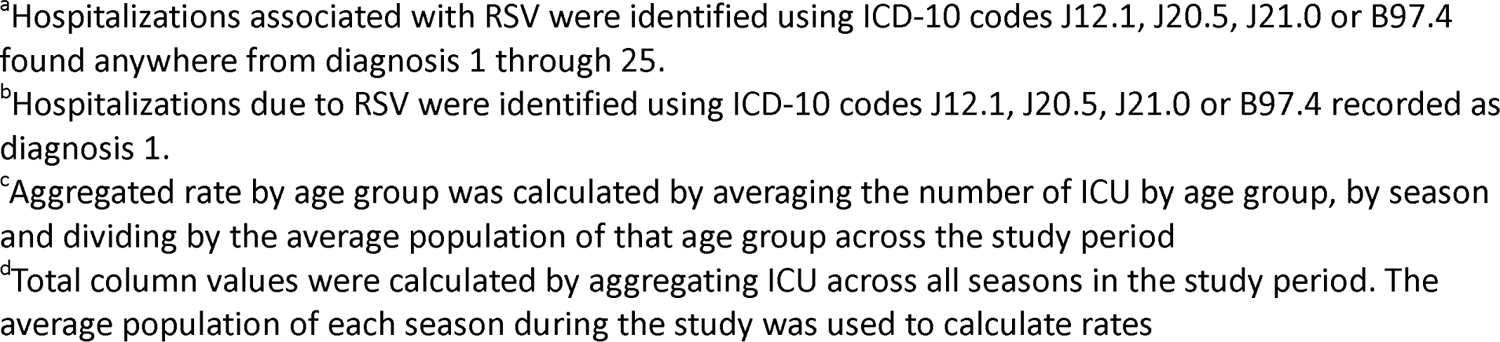
Number and rate of ICU admissions associated with and due to RSV, percentage of RSV hospitalizations resulting in an ICU admission, by age group, seasons 2010-2011 to 2019-2020 and 2021-2022 to 2022-2023, Canada (excluding Quebec), Canadian Discharge Abstract Database.

| **Age Group** | **Among hospitalizations associated with RSV<sup>a</sup>** |  |  | **Among hospitalizations due to RSV<sup>b</sup>** |  |  |
| --- | --- | --- | --- | --- | --- | --- |
|  | **Number of ICU admissions** | **Rate per 100,000 population<sup>c</sup>** | **% of hospitalizations requiring ICU** | **Number of ICU admissions** | **Rate per 100,000 population<sup>c</sup>** | **% of hospitalizations requiring ICU** |
| 50-59 | 504 | 1.1 | 26 | 101 | 0.2 | 17 |
| 60-69 | 896 | 2.4 | 23 | 155 | 0.4 | 14 |
| 70-79 | 969 | 4.3 | 19 | 174 | 0.8 | 12 |
| 80+ | 815 | 6.0 | 9 | 212 | 1.5 | 7 |
| **Total<sup>d</sup>** | **3,184** | **2.6** | **16%** | **642** | **0.5** | **10%** |

<sup>a</sup>Hospitalizations associated with RSV were identified using ICD-10 codes J12.1, J20.5, J21.0 or B97.4 found anywhere from diagnosis 1 through 25.
<sup>b</sup>Hospitalizations due to RSV were identified using ICD-10 codes J12.1, J20.5, J21.0 or B97.4 recorded as diagnosis 1.
<sup>c</sup>Aggregated rate by age group was calculated by averaging the number of ICU by age group, by season and dividing by the average population of that age group across the study period
<sup>d</sup>Total column values were calculated by aggregating ICU across all seasons in the study period. The average population of each season during the study was used to calculate rates

Regardless of the type of RSV hospitalization (associated with or due to), the number and rate of ICU admissions increased with increasing age but the proportion of hospitalizations requiring ICU admissions decreased with increasing age.

### Death associated with RSV Infection

#### Rapid Review

Five SRs and eleven observational studies, including four Canadian studies, reported data on death associated with RSV infection. Although evidence is more limited than for other clinical outcomes, in general the CFR among adults admitted to hospital is approximately 5-10% which increases with increasing age and the presence of one or more comorbidities. A SR of developed countries found an overall RSV-related case fatality ratio (CFR) of 8.2% (95% CI: 5.5-11.9%) among adults 60 years of age and older and 9.9% (95% CI: 6.7-14.4%) among adults 18 years of age and older considered at higher risk (7). Another systematic review and meta-analysis found that the in-hospital case fatality rate was higher in adults 65 years of age and older than adults 50 to 64 years of age (1). Similarly, two studies from Ontario found that among patients hospitalized with RSV, 30-day all-cause mortality rates increased with increasing age (12, 17). A US prospective cohort study found that the CFR was higher in adults admitted from LTC facilities (38%) than in those admitted from the community (3%, p<0.001) (18).

#### Canadian Hospitalization Data

Across 12 seasons, 1,668 in-hospital deaths among RSV associated hospitalizations were reported in adults 50 years of age and older, corresponding to an in-hospital CFR of 9% (Table 5). Overall, 397 of these in-hospital deaths were among those hospitalized due to RSV, corresponding to an in-hospital CFR of 6%. The average rate of in-hospital deaths associated with RSV in adults 50 years of age and older across the 12 seasons was 1.4 per 100,000 population and increased with increasing age (the rates of in-hospital deaths in hospitalizations associated with RSV per 100,000 population by age groups were the following: 0.2 in adults 50 to 59 years old, 0.6 in 60 to 69 years, 1.7 in 70 to 79 years, and 6.7 in adults 80 years of age and older). The rates of in-hospital deaths among older adults hospitalized due to RSV in Canada followed the same trend; however, rates were much lower. Regardless of the type of RSV death (in hospitalizations associated with or due to RSV), both the number and rates of death and CFR increased with increasing age.

**Table 5:** Number and rate of in-hospital deaths associated with and due to RSV, case fatality rate (CFR), by age group, seasons 2010-2011 to 2019-2020 and 2021-2022 to 2022-2023, Canada (excluding Quebec), Canadian Discharge Abstract Database.

| Age Group | Among hospitalizations associated with RSV<sup>a</sup> |  |  | Among hospitalizations due to RSV<sup>b</sup> |  |  |
| --- | --- | --- | --- | --- | --- | --- |
|  | Number of in-hospital deaths | Rate per 100,000 population<sup>c</sup> | CFR (%) | Number of in-hospital deaths | Rate per 100,000 population<sup>c</sup> | CFR (%) |
| 50-59 | 110 | 0.2 | 6 | 18 | 0.0 | 3 |
| 60-69 | 244 | 0.6 | 6 | 44 | 0.1 | 4 |
| 70-79 | 390 | 1.7 | 8 | 72 | 0.3 | 5 |
| 80+ | 924 | 6.7 | 11 | 263 | 1.9 | 8 |
| **Total<sup>d</sup>** | **1668** | **1.4** | **9** | **397** | **0.3** | **6** |

<sup>a</sup>Hospitalizations associated with RSV were identified using ICD-10 codes J12.1, J20.5, J21.0 or B97.4 found anywhere from diagnosis 1 through 25.
<sup>b</sup>Hospitalizations due to RSV were identified using ICD-10 codes J12.1, J20.5, J21.0 or B97.4 recorded as diagnosis 1.
<sup>c</sup>Aggregated rate by age group was calculated by averaging the number of deaths by age group, by season and dividing by the average population of that age group across the study period
<sup>d</sup>Total column values were calculated by aggregating deaths across all seasons in the study period. The average population of each season during the study was used to calculate rates.

## Discussion

The rapid review offers insight into the burden of RSV disease in older adults and adults with underlying medical conditions, with a focus on high-income countries such as Canada, the US and European countries. This review is also supported with hospitalization data to further describe RSV burden of disease in Canada.

Evidence from the rapid review suggest that medically attended RSV infections in high-income countries are frequent in older adults and those with underlying medical conditions. The incidence of RSV RTI increases with increasing age as well as the presence of comorbidities, including cardiorespiratory disease, diabetes and immunocompromising conditions. While the incidence of hospitalization varies between studies, risk of hospitalization associated with RSV increases consistently with increasing age. Depending on age and risk factors, adults 18 years of age and older with underlying medical conditions are more likely to have a hospitalization associated with RSV infection than those without. Patients who reside in LTC or other chronic care facilities have a higher likelihood of severe clinical outcomes compared to patients with other living situations upon hospital admission. Moreover, ICU admission associated with RSV increases with increasing age and presence of comorbidities, with approximately 10% of older hospitalized older adults requiring ICU admission. There were more limited data on burden of death associated with RSV. The CFR among those admitted to hospital varied between studies but is approximately 5 to 10% and increases with increasing age.

Canadian administrative hospitalization data generally support the findings of the rapid review. Over 12 respiratory seasons between August 2010 and September 2023, it was found that RSV-associated hospitalization rates increased with increasing age and that finding was consistent for each season. The average rates of hospitalization associated with RSV in adults 50 years of age and older was estimated at 16 per 100,000. Overall, 16% of hospitalizations associated with RSV resulted in an ICU admission corresponding to an average rate of 2.6 per 100,000 for adults 50 years of age and older. Rate of hospitalization associated with RSV resulting in ICU admissions increased with increasing age; however, the proportion of hospitalizations requiring ICU admission decreased with increasing age. The average CFR among adults 50 years of age and older was 9% and in-hospital death among hospitalizations associated with RSV increased with increasing age. Hospitalizations, ICU admissions and deaths due to RSV followed the same trend; however, calculated values were lower than those associated with RSV.

Although there is general alignment between the rapid review and Canadian hospitalization data analysis, some differences can be noted. Incidence rates derived from the Canadian hospitalization data were usually lower than what is reported in the literature. Discrepancies can be explained by differences in methodology used between studies. Individual study characteristics such as study population, case definitions, study period, and data source can have an important impact on study results generalizability; thus, leading to discrepancies between the observed incidence rates. As for the rapid review, it included several studies, such as systematic reviews and meta-analysis, as well as large prospective population-based cohort studies, which represent a comprehensive picture of the current literature on RSV burden of disease in adults. However, few studies reported data on Canadian adults and heterogeneity between study results limits the generalizability of the findings. Another limitation of the rapid review is the inclusion of RSV infection not limited to laboratory confirmed infection potentially leading to an overestimation of RSV incidence.

Currently, Canada has limited enhanced national RSV surveillance data and leveraging administrative health data from CIHI DAD helped address those evidence gaps to supplement the evidence on RSV burden of disease in adults to inform the development of immunization recommendation. However, known limitations of healthcare administrative data are expected to lead to underestimation of RSV incidence especially due to limits in viral identification and undertesting in patients (19). Of note, rates from CIHI DAD were higher in the more recent period, which could be partly due to more frequent testing Other limitations of the Canadian hospitalization data include the exclusion of Quebec data, a large Canadian province, and changes in practice during the study period, especially in respiratory season following the COVID-19 pandemic.

The descriptive analyses provided information on general trends of severe outcomes of RSV RTI in older adults (19). Although the rapid review and healthcare administrative data analysis methodologies each have their drawbacks, the combination of these analyses provides an interdisciplinary view of the burden of RSV in older adults to support vaccine program decision making. Enhanced National surveillance programs for RSV are in development where timely data variables of interest can be collected specifically for surveillance activities and to support policy and decision making. These analyses may be revisited as additional data becomes available from the literature or from the Canadian surveillance landscape.

## Data Availability

All data produced in the present work are contained in the manuscript

## Acknowledgements

The authors wish to acknowledge the NACI Secretariat team, Matthew Tunis, Kelsey Young, Mona Hersi, Adrienne Stevens, Anastassia Howarth, Su Hyun Lim, the Health Canada Library (Shannon Hayes) and the NACI RSV Working Group.

## Author’s Statement

EA: conceptualization, analysis and interpretation of data, drafting paper

PDP: analysis and interpretation of data, drafting the paper

PD: analysis and interpretation of data, revising the paper

AR: analysis and interpretation of the data

LL: interpretation of the data and drafting the paper

NB: revising the paper

WS: conceptualization, revising the paper

AK: conceptualization, drafting paper

## Notes

### Competing Interest Statement

The authors have declared no competing interest.

### Funding Statement

This study did not receive any funding

## References

1. Shi T, Denouel A, Tietjen AK, Campbell I, Moran E, Li X, et al. Global disease burden estimates of respiratory syncytial virus-associated acute respiratory infection in older adults in 2015: A systematic review and meta-analysis. J Infect Dis. 2020 Oct 07;222(Suppl 7):S577–83. 10.1093/infdis/jiz059.

2. ElSherif M, Andrew MK, Ye L, Ambrose A, Boivin G, Bowie W, et al. Leveraging influenza virus surveillance from 2012 to 2015 to characterize the burden of respiratory syncytial virus disease in Canadian adults ≥50 years of age hospitalized with acute respiratory illness. Open Forum Infect Dis. 2023 Jun 13;10(7):ofad315. 10.1093/ofid/ofad315.

3. Mesa-Frias M, Rossi C, Emond B, Bookhart B, Anderson D, Drummond S, et al. Incidence and economic burden of respiratory syncytial virus among adults in the United States: A retrospective analysis using 2 insurance claims databases. J Manag Care Spec Pharm. 2022 Jul;28(7):753–65. 10.18553/jmcp.2022.21459.

4. Discharge Abstract Database (DAD) metadata [Internet]. Canadian Institute for Health Information; 2023 [cited 2023 Nov 27]. Available from: https://www.cihi.ca/en/discharge-abstract-database-dad-metadata.

5. Statistics Canada. Table 17-10-0005-01 Population estimates on July 1, by age and gender [Internet]. Ottawa (ON): Government of Canada; 2024 [cited 2024 Feb 16]. Available from: 10.25318/1710000501-eng.

6. Public Health Agency of Canada (PHAC). Respiratory virus report, week 34 - ending August 28, 2021 [Internet]. Ottawa (ON): Government of Canada; 2021 Sep 02 [cited 2024 Feb 16]. Available from: https://www.canada.ca/en/public-health/services/surveillance/respiratory-virus-detections-canada/2021-2022/week-34-ending-august-28-2021.html.

7. Nguyen-Van-Tam JS, O’Leary M, Martin ET, Heijnen E, Callendret B, Fleischhackl R, et al. Burden of respiratory syncytial virus infection in older and high-risk adults: A systematic review and meta-analysis of the evidence from developed countries. Eur Respir Rev. 2022 Dec;31(166):220105. 10.1183/16000617.0105-2022.

8. Wilkinson T, Beaver S, Macartney M, McArthur E, Yadav V, Lied-Lied A. Burden of respiratory syncytial virus in adults in the United Kingdom: A systematic literature review and gap analysis. Influenza Other Respir Viruses. 2023 Sep;17(9):e13188. 10.1111/irv.13188.

9. Johannesen CK, van Wijhe M, Tong S, Fernández LV, Heikkinen T, van Boven M, et al. Age-specific estimates of respiratory syncytial virus-associated hospitalizations in 6 European countries: A time series analysis. J Infect Dis. 2022 Aug 12;226(Suppl 1):S29–37. 10.1093/infdis/jiac150.

10. McLaughlin JM, Khan F, Begier E, Swerdlow DL, Jodar L, Falsey AR. Rates of medically attended RSV among US adults: A systematic review and meta-analysis. Open Forum Infect Dis. 2022 Jul;9(7):ofac300. 10.1093/ofid/ofac300.

11. Belongia EA, King JP, Kieke BA, Pluta J, Al-Hilli A, Meece JK, et al. Clinical features, severity, and incidence of RSV illness during 12 consecutive seasons in a community cohort of adults ≥60 years old. Open Forum Infect Dis. 2018 Dec;5(12):ofy316. 10.1093/ofid/ofy316.

12. Mac S, Shi S, Millson B, Tehrani A, Eberg M, Myageri V, et al. Burden of illness associated with Respiratory Syncytial Virus (RSV)-related hospitalizations among adults in Ontario, Canada: A retrospective population-based study. Vaccine. 2023 Aug 07;41(35):5141–9. 10.1016/j.vaccine.2023.06.071.

13. Public Health Agency of Canada (PHAC). Respiratory virus report, week 34 - ending August 27, 2022 [Internet]. Ottawa (ON): Government of Canada; 2022 Sep 01 [cited 2024 Jan 17]. Available from: https://www.canada.ca/en/public-health/services/surveillance/respiratory-virus-detections-canada/2021-2022/week-34-ending-august-27-2022.html.

14. Colosia AD, Yang J, Hillson E, Mauskopf J, Copley-Merriman C, Shinde V, et al. The epidemiology of medically attended respiratory syncytial virus in older adults in the United States: A systematic review. PLoS One. 2017;12(8):e0182321. 10.1371/journal.pone.0182321.

15. Mulpuru S, Andrew MK, Ye L, Hatchette T, LeBlanc J, El-Sherif M, et al. Impact of respiratory viral infections on mortality and critical illness among hospitalized patients with chronic obstructive pulmonary disease. Influenza Other Respir Viruses. 2022 Nov;16(6):1172–82. 10.1111/irv.13050.

16. Goldman CR, Sieling WD, Alba LR, Silverio Francisco RA, Vargas CY, Barrett AE, et al. Severe clinical outcomes among adults hospitalized with respiratory syncytial virus infections, New York City, 2017-2019. Public Health Rep. 2022 Sep;137(5):929–35. 10.1177/00333549211041545.

17. Hamilton MA, Liu Y, Calzavara A, Sundaram ME, Djebli M, Darvin D, et al. Predictors of all-cause mortality among patients hospitalized with influenza, respiratory syncytial virus, or SARS-CoV-2. Influenza Other Respir Viruses. 2022 Nov;16(6):1072–81. 10.1111/irv.13004.

18. Falsey AR, Hennessey PA, Formica MA, Cox C, Walsh EE. Respiratory syncytial virus infection in elderly and high-risk adults. N Engl J Med. 2005 Apr 28;352(17):1749–59. 10.1056/NEJMoa043951.

19. Schanzer DL, Saboui M, Lee L, Nwosu A, Bancej C. Burden of influenza, respiratory syncytial virus, and other respiratory viruses and the completeness of respiratory viral identification among respiratory inpatients, Canada, 2003-2014. Influenza Other Respir Viruses. 2018 Jan;12(1):113–21. 10.1111/irv.12497.

20. Savic M, Penders Y, Shi T, Branche A, Pirçon J. Respiratory syncytial virus disease burden in adults aged 601years and older in high-income countries: A systematic literature review and meta-analysis. Influenza Other Respir Viruses. 2023 Jan;17(1):e13031. 10.1111/irv.13031.

21. Shi T, Vennard S, Jasiewicz F, Brogden R, Nair H. Disease Burden Estimates of Respiratory Syncytial Virus related Acute Respiratory Infections in Adults With Comorbidity: A Systematic Review and Meta-Analysis. J Infect Dis. 2022 Aug 12;226(Suppl 1):S17–21. 10.1093/infdis/jiab040.

22. Wyffels V, Kariburyo F, Gavart S, Fleischhackl R, Yuce H. A real-world analysis of patient characteristics and predictors of hospitalization among US medicare beneficiaries with respiratory syncytial virus infection. Adv Ther. 2020 Mar;37(3):1203–17. 10.1007/s12325-020-01230-3.

23. Falsey AR, McElhaney JE, Beran J, van Essen GA, Duval X, Esen M, et al. Respiratory syncytial virus and other respiratory viral infections in older adults with moderate to severe influenza-like illness. J Infect Dis. 2014 Jun 15;209(12):1873–81. 10.1093/infdis/jit839.

24. Sundaram ME, Meece JK, Sifakis F, Gasser RA, Belongia EA. Medically attended respiratory syncytial virus infections in adults aged ≥ 50 years: Clinical characteristics and outcomes. Clin Infect Dis. 2014 Feb;58(3):342–9. 10.1093/cid/cit767.

25. De Serres G, Lampron N, La Forge J, Rouleau I, Bourbeau J, Weiss K, et al. Importance of viral and bacterial infections in chronic obstructive pulmonary disease exacerbations. J Clin Virol. 2009 Oct;46(2):129–33. 10.1016/j.jcv.2009.07.010.

26. Schanzer DL, Langley JM, Tam TWS. Role of influenza and other respiratory viruses in admissions of adults to Canadian hospitals. Influenza Other Respir Viruses. 2008 Jan;2(1):1–8. 10.1111/j.1750-2659.2008.00035.x.

27. Havers FP, Whitaker M, Melgar M, Chatwani B, Chai SJ, Alden NB, et al. Characteristics and outcomes among adults aged ≥60 years hospitalized with laboratory-confirmed respiratory syncytial virus - RSV-NET, 12 states, July 2022-June 2023. MMWR Morb Mortal Wkly Rep. 2023 Oct 06;72(40):1075–82. 10.15585/mmwr.mm7240a1.

28. Branche AR, Saiman L, Walsh EE, Falsey AR, Sieling WD, Greendyke W, et al. Incidence of respiratory syncytial virus infection among hospitalized adults, 2017-2020. Clin Infect Dis. 2022 Mar 23;74(6):1004–11. 10.1093/cid/ciab595.

29. Zheng Z, Warren JL, Shapiro ED, Pitzer VE, Weinberger DM. Estimated incidence of respiratory hospitalizations attributable to RSV infections across age and socioeconomic groups. Pneumonia (Nathan). 2022 Oct 25;14(1):6. 10.1186/s41479-022-00098-x.

30. Walsh E, Lee N, Sander I, Stolper R, Zakar J, Wyffels V, et al. RSV-associated hospitalization in adults in the USA: A retrospective chart review investigating burden, management strategies, and outcomes. Health Sci Rep. 2022 Apr;5(3):e556. 10.1002/hsr2.556.

31. Ellis SE, Coffey CS, Mitchel EF, Dittus RS, Griffin MR. Influenza- and respiratory syncytial virus-associated morbidity and mortality in the nursing home population. J Am Geriatr Soc. 2003 Jun;51(6):761–7. 10.1046/j.1365-2389.2003.51254.x.

32. Tseng HF, Sy LS, Ackerson B, Solano Z, Slezak J, Luo Y, et al. Severe morbidity and short- and mid- to long-term mortality in older adults hospitalized with respiratory syncytial virus infection. J Infect Dis. 2020 Sep 14;222(8):1298–310. 10.1093/infdis/jiaa361.

