## Supplementary material for "Burden of Disease of Respiratory Syncytial Virus in Older Adults and Adults Considered at High Risk of Severe Infection": CCDR older adult_Supplementary material.docx

Elissa Abrams, MD MPH FRCPC

Pamela Doyon-Plourde, PhD

Phaedra Davis, MPH

Liza Lee, MSc

Abbas Rahal, PhD

Nicholas Brousseau, MD

Winnie Siu, MD MSc CCFP FRCPC

April Killikelly, PhD

### Supplementary material S1

Search strategy for studies evaluating the burden of respiratory syncytial virus infection and its complications in adults.

#### Table S1: Search strategy for Medline database(s), Ovid MEDLINE(R) ALL 1946 to November 11, 2022

| **#** | **Searches** | **Results** |
| --- | --- | --- |
| 1 | Respiratory Syncytial Virus Infections/ | 8320 |
| 2 | (respiratory syncytial vir* or RSV or hrsv).ti,kf,kw. | 11134 |
| 3 | (respiratory syncytial and (rsv or hrsv)).ab. | 9389 |
| 4 | 1 or 2 or 3 [RSV] | 14414 |
| 5 | exp adult/ | 7860444 |
| 6 | (adult? or women or men or man or woman or senior? or elderly or ((old or older) adj2 (patient* or inpatient* or outpatient*))).tw,kf,kw. | 3521304 |
| 7 | ((aged or ages or year?) adj2 ("18" or "19" or "2#" or "3#" or "4#" or "5#" or "6#" or "7#" or "8#" or "9#")).tw,kf. | 2141470 |
| 8 | or/5-7 [adults] | 9723524 |
| 9 | 4 and 8 [RSV + adults] | 3246 |
| 10 | (meta-analysis or systematic review).pt. | 290969 |
| 11 | meta-analysis/ or systematic review/ or meta-analysis as topic/ or systematic review as topic/ | 315543 |
| 12 | ((systematic* adj3 (review* or overview*)) or (methodologic* adj3 (review* or overview*))).ti,ab,kf. | 291527 |
| 13 | ((quantitative adj3 (review* or overview* or synthes*)) or (research adj3 (integrati* or overview*))).ti,ab,kf. | 14572 |
| 14 | (meta analy* or metanaly*).ti,ab,kf,kw. | 251865 |
| 15 | (cochrane or evidence report).jw. | 16353 |
| 16 | (comparative adj3 (efficacy or effectiveness)).ti,ab,kf. | 16675 |
| 17 | or/10-16 [adapted from "SR / MA / HTA / ITC - MEDLINE, Embase, PsycInfo. In: CADTH Search Filters Database. Ottawa: CADTH; 2022: <https://searchfilters.cadth.ca/link/33>." Accessed 2022-10-28.] | 469518 |
| 18 | 9 and 17 | 63 |
| 19 | limit 18 to yr="1995-current" | 63 |
| 20 | risk factor/ or Social Determinants of Health/ or exp Health Inequities/ | 957378 |
| 21 | (determinant* or risk or risks or probabilit* or probable or susceptib* or vulnerab* or inequalit* or inequit* or likely or predisposed or likelihood).ti,kf,kw. | 904232 |
| 22 | (determinant* or risk or risks or probabilit* or probable or susceptib* or vulnerab* or inequalit* or inequit* or likely or predisposed or likelihood).ab. /freq=2 | 1811585 |
| 23 | 20 or 21 or 22 | 2592306 |
| 24 | 9 and 23 | 571 |
| 25 | (case reports or comment or editorial or letter).pt. | 4183845 |
| 26 | 24 not 25 | 562 |
| 27 | limit 26 to yr="1995-current" | 546 |

#### Table S2: Search strategy for Embase database(s), Embase 1974 to November 11, 2022

| **#** | **Searches** | **Results** |
| --- | --- | --- |
| 1 | exp *Human respiratory syncytial virus/ | 2139 |
| 2 | (respiratory syncytial vir* or RSV or hrsv).ti,kf,kw. | 13781 |
| 3 | (respiratory syncytial and (rsv or hrsv)).ab. | 11662 |
| 4 | 1 or 2 or 3 [RSV] | 16703 |
| 5 | Adult/ or middle aged/ or young adult/ or exp aged/ | 10099351 |
| 6 | (adult? or women or men or man or woman or senior? or elderly or ((old or older) adj2 (patient* or inpatient* or outpatient*))).tw,kf,kw. | 4743887 |
| 7 | ((aged or ages or year?) adj2 ("18" or "19" or "2#" or "3#" or "4#" or "5#" or "6#" or "7#" or "8#" or "9#")).tw,kf. | 3354668 |
| 8 | or/5-7 [adults] | 12461878 |
| 9 | 4 and 8 [RSV + adults] | 3995 |
| 10 | meta-analysis/ or systematic review/ or "meta analysis (topic)"/ or "systematic review (topic)"/ | 553065 |
| 11 | ((systematic* adj3 (review* or overview*)) or (methodologic* adj3 (review* or overview*))).ti,ab,kf. | 356521 |
| 12 | ((quantitative adj3 (review* or overview* or synthes*)) or (research adj3 (integrati* or overview*))).ti,ab,kf. | 16983 |
| 13 | (meta analy* or metanaly*).ti,ab,kf. | 320658 |
| 14 | (cochrane or evidence report).jw. | 24057 |
| 15 | (comparative adj3 (efficacy or effectiveness)).ti,ab,kf. | 24343 |
| 16 | or/10-15 [adapted from "SR / MA / HTA / ITC - MEDLINE, Embase, PsycInfo. In: CADTH Search Filters Database. Ottawa: CADTH; 2022: <https://searchfilters.cadth.ca/link/33>." Accessed 2022-10-28.] | 714515 |
| 17 | 9 and 16 | 101 |
| 18 | limit 17 to yr="1995-current" | 101 |
| 19 | *risk factor/ or *"social determinants of health"/ or *health disparity/ | 128595 |
| 20 | (determinant* or risk or risks or probabilit* or probable or susceptib* or vulnerab* or inequalit* or inequit* or likely or predisposed or likelihood).ti,kf,kw. | 1223779 |
| 21 | (determinant* or risk or risks or probabilit* or probable or susceptib* or vulnerab* or inequalit* or inequit* or likely or predisposed or likelihood).ab. /freq=2 | 2576907 |
| 22 | or/19-21 | 3062321 |
| 23 | 9 and 22 | 700 |
| 24 | (letter or note or editorial).pt. | 2901556 |
| 25 | case report?.ti. | 378001 |
| 26 | 23 not (24 or 25) | 696 |
| 27 | limit 26 to yr="1995-current" | 683 |

#### Table S3: Search strategy for Global Health database(s), Global Health 1973 to 2022 Week 45

| **#** | **Searches** | **Results** |
| --- | --- | --- |
| 1 | Human respiratory syncytial virus/ | 6751 |
| 2 | (respiratory syncytial vir* or RSV or hrsv).ti,hw,id. | 7179 |
| 3 | (respiratory syncytial and (rsv or hrsv)).ab. | 4648 |
| 4 | 1 or 2 or 3 [RSV] | 7367 |
| 5 | adults/ | 87624 |
| 6 | (adult? or women or men or man or woman or senior? or elderly or ((old or older) adj2 (patient* or inpatient* or outpatient*))).ti,ab. | 723233 |
| 7 | (adult? or women or men or woman or senior? or elderly or ((old or older) adj2 (patient* or inpatient* or outpatient*))).hw,id. | 475302 |
| 8 | ((aged or ages or year?) adj2 ("18" or "19" or "2#" or "3#" or "4#" or "5#" or "6#" or "7#" or "8#" or "9#")).ti,ab. | 410496 |
| 9 | or/5-8 [adults] | 978885 |
| 10 | 4 and 9 [RSV + adults] | 1738 |
| 11 | meta-analysis/ or systematic reviews/ | 69684 |
| 12 | ((systematic* adj3 (review* or overview*)) or (methodologic* adj3 (review* or overview*))).ti,ab,hw. | 64136 |
| 13 | ((quantitative adj3 (review* or overview* or synthes*)) or (research adj3 (integrati* or overview*))).ti,ab,hw. | 2487 |
| 14 | (meta analy* or metanaly*).ti,ab,hw,id. | 50746 |
| 15 | (cochrane or evidence report).jx. | 951 |
| 16 | (comparative adj3 (efficacy or effectiveness)).ti,ab,hw. | 2191 |
| 17 | or/11-16 [adapted from "SR / MA / HTA / ITC - MEDLINE, Embase, PsycInfo. In: CADTH Search Filters Database. Ottawa: CADTH; 2022: <https://searchfilters.cadth.ca/link/33>." Accessed 2022-10-28.] | 87577 |
| 18 | 10 and 17 | 55 |
| 19 | limit 18 to yr="1995-current" | 55 |
| 20 | risk factor/ or health inequalities/ | 320571 |
| 21 | (determinant* or risk or risks or probabilit* or probable or susceptib* or vulnerab* or inequalit* or inequit* or likely or predisposed or likelihood).ti. | 246011 |
| 22 | (determinant* or risk or risks or probabilit* or probable or susceptib* or vulnerab* or inequalit* or inequit* or likely or predisposed or likelihood).ab. /freq=2 | 506457 |
| 23 | or/20-22 | 678737 |
| 24 | 10 and 23 | 374 |
| 25 | editorial.pt. | 3030 |
| 26 | case report?.ti. | 18723 |
| 27 | 24 not (25 or 26) | 374 |
| 28 | limit 27 to yr="1995-current" | 371 |

#### Search strategy for ProQuest Public Health limited to systematic review: 12 results

(((TITLE(respiratory syncytial vir* or RSV or hrsv) OR SUBJECT(respiratory syncytial vir* or RSV or hrsv) OR MJMESH( Respiratory Syncytial Virus Infections)) OR ABSTRACT(respiratory syncytial AND (rsv OR hrsv))) AND (MESH(adult) OR TITLE(adult or adults or women or men or man or woman or senior or seniors or elderly or ((old or older) NEAR/2 (patient* or inpatient* or outpatient*))) OR ABSTRACT(adult or adults or women or men or man or woman or senior or seniors or elderly or ((old or older) NEAR/2 (patient* or inpatient* or outpatient*))))) AND (MESH(meta-analysis) or MESH(systematic review) or MESH(meta-analysis as topic) or MESH(systematic review as topic) OR TITLE((systematic* NEAR/3 (review* or overview*)) or (methodologic* NEAR/3 (review* or overview*))) OR ABSTRACT((systematic* NEAR/3 (review* or overview*)) or (methodologic* NEAR/3 (review* or overview*))) OR TITLE((quantitative NEAR/3 (review* or overview* or synthes*)) or (research NEAR/3 (integrati* or overview*))) OR ABSTRACT((quantitative NEAR/3 (review* or overview* or synthes*)) or (research NEAR/3 (integrati* or overview*))) OR TITLE(meta analy* or metanaly*) OR ABSTRACT(meta analy* or metanaly*) OR TITLE(comparative NEAR/3 (efficacy or effectiveness)) OR ABSTRACT(comparative NEAR/3 (efficacy or effectiveness))) AND pd(19950101-20231231)

#### Search strategy for ProQuest Public Health: 212 results

(((TITLE(respiratory syncytial vir* or RSV or hrsv) OR SUBJECT(respiratory syncytial vir* or RSV or hrsv) OR MJMESH( Respiratory Syncytial Virus Infections)) OR ABSTRACT(respiratory syncytial AND (rsv OR hrsv))) AND (MESH(adult) OR TITLE(adult or adults or women or men or man or woman or senior or seniors or elderly or ((old or older) NEAR/2 (patient* or inpatient* or outpatient*))) OR ABSTRACT(adult or adults or women or men or man or woman or senior or seniors or elderly or ((old or older) NEAR/2 (patient* or inpatient* or outpatient*))))) AND (MESH(risk factor) OR MESH(Social Determinants of Health) OR MESH(Health Inequities) OR TITLE(determinant* or risk or risks or probabilit* or probable or susceptib* or vulnerab* or inequalit* or inequit* or likely or predisposed or likelihood) OR ABSTRACT(determinant* or risk or risks or probabilit* or probable or susceptib* or vulnerab* or inequalit* or inequit* or likely or predisposed or likelihood)) AND pd(19950101-20231231)

### Supplementary material S2

Search strategy update for studies evaluating the burden of respiratory syncytial virus infection and its complications in adults.

#### Table S4: Search strategy for Medline database(s), Ovid MEDLINE(R) ALL 1946 to September 27, 2023

| **#** | **Searches** | **Results** |
| --- | --- | --- |
| 1 | Respiratory Syncytial Virus Infections/ | 8769 |
| 2 | (respiratory syncytial vir* or RSV or hrsv).ti,kf,kw. | 11914 |
| 3 | (respiratory syncytial and (rsv or hrsv)).ab. | 10048 |
| 4 | 1 or 2 or 3 [RSV] | 15348 |
| 5 | exp adult/ | 7968450 |
| 6 | (adult? or women or men or man or woman or senior? or elderly or ((old or older) adj2 (patient* or inpatient* or outpatient*))).tw,kf,kw. | 3699303 |
| 7 | ((aged or ages or year?) adj2 ("18" or "19" or "2#" or "3#" or "4#" or "5#" or "6#" or "7#" or "8#" or "9#")).tw,kf. | 2267222 |
| 8 | or/5-7 [adults] | 9977878 |
| 9 | 4 and 8 [RSV + adults] | 3511 |
| 10 | (meta-analysis or systematic review).pt. | 323489 |
| 11 | meta-analysis/ or systematic review/ or meta-analysis as topic/ or systematic review as topic/ | 348395 |
| 12 | ((systematic* adj3 (review* or overview*)) or (methodologic* adj3 (review* or overview*))).ti,ab,kf. | 331840 |
| 13 | ((quantitative adj3 (review* or overview* or synthes*)) or (research adj3 (integrati* or overview*))).ti,ab,kf. | 16068 |
| 14 | (meta analy* or metanaly*).ti,ab,kf,kw. | 282356 |
| 15 | (cochrane or evidence report).jw. | 16760 |
| 16 | (comparative adj3 (efficacy or effectiveness)).ti,ab,kf. | 17888 |
| 17 | or/10-16 [adapted from "SR / MA / HTA / ITC - MEDLINE, Embase, PsycInfo. In: CADTH Search Filters Database. Ottawa: CADTH; 2022: <https://searchfilters.cadth.ca/link/33>." Accessed 2022-10-28.] | 520845 |
| 18 | 9 and 17 | 80 |
| 19 | limit 18 to yr="1995-current" | 80 |
| 20 | risk factor/ or Social Determinants of Health/ or exp Health Inequities/ | 983278 |
| 21 | (determinant* or risk or risks or probabilit* or probable or susceptib* or vulnerab* or inequalit* or inequit* or likely or predisposed or likelihood).ti,kf,kw. | 968595 |
| 22 | (determinant* or risk or risks or probabilit* or probable or susceptib* or vulnerab* or inequalit* or inequit* or likely or predisposed or likelihood).ab. /freq=2 | 1942047 |
| 23 | 20 or 21 or 22 | 2750919 |
| 24 | 9 and 23 | 614 |
| 25 | (case reports or comment or editorial or letter).pt. | 4322092 |
| 26 | 24 not 25 | 605 |
| 27 | ("20221113" or "20221114" or "20221115" or "20221116" or "20221117" or "20221118" or "20221119" or 2022112* or 2022113* or 202212* or 2023*).dt,dp. | 1514878 |
| 28 | 26 and 27 | 45 |

#### Table S5: Search strategy for Embase database(s), Embase 1974 to September 27, 2023

| **#** | **Searches** | **Results** |
| --- | --- | --- |
| 1 | exp *Human respiratory syncytial virus/ | 2506 |
| 2 | (respiratory syncytial vir* or RSV or hrsv).ti,kf,kw. | 14802 |
| 3 | (respiratory syncytial and (rsv or hrsv)).ab. | 12537 |
| 4 | 1 or 2 or 3 [RSV] | 17916 |
| 5 | Adult/ or middle aged/ or young adult/ or exp aged/ | 10760904 |
| 6 | (adult? or women or men or man or woman or senior? or elderly or ((old or older) adj2 (patient* or inpatient* or outpatient*))).tw,kf,kw. | 5012431 |
| 7 | ((aged or ages or year?) adj2 ("18" or "19" or "2#" or "3#" or "4#" or "5#" or "6#" or "7#" or "8#" or "9#")).tw,kf. | 3571575 |
| 8 | or/5-7 [adults] | 13205102 |
| 9 | 4 and 8 [RSV + adults] | 4431 |
| 10 | meta-analysis/ or systematic review/ or "meta analysis (topic)"/ or "systematic review (topic)"/ | 617899 |
| 11 | ((systematic* adj3 (review* or overview*)) or (methodologic* adj3 (review* or overview*))).ti,ab,kf. | 403915 |
| 12 | ((quantitative adj3 (review* or overview* or synthes*)) or (research adj3 (integrati* or overview*))).ti,ab,kf. | 18643 |
| 13 | (meta analy* or metanaly*).ti,ab,kf. | 357755 |
| 14 | (cochrane or evidence report).jw. | 25059 |
| 15 | (comparative adj3 (efficacy or effectiveness)).ti,ab,kf. | 26188 |
| 16 | or/10-15 [adapted from "SR / MA / HTA / ITC - MEDLINE, Embase, PsycInfo. In: CADTH Search Filters Database. Ottawa: CADTH; 2022: <https://searchfilters.cadth.ca/link/33>." Accessed 2022-10-28.] | 792704 |
| 17 | 9 and 16 | 133 |
| 18 | limit 17 to yr="1995-current" | 133 |
| 19 | *risk factor/ or *"social determinants of health"/ or *health disparity/ | 144981 |
| 20 | (determinant* or risk or risks or probabilit* or probable or susceptib* or vulnerab* or inequalit* or inequit* or likely or predisposed or likelihood).ti,kf,kw. | 1309513 |
| 21 | (determinant* or risk or risks or probabilit* or probable or susceptib* or vulnerab* or inequalit* or inequit* or likely or predisposed or likelihood).ab. /freq=2 | 2768741 |
| 22 | or/19-21 | 3283728 |
| 23 | 9 and 22 | 779 |
| 24 | (letter or note or editorial).pt. | 3029967 |
| 25 | case report?.ti. | 406269 |
| 26 | 23 not (24 or 25) | 775 |
| 27 | ("20221113" or "20221114" or "20221115" or "20221116" or "20221117" or "20221118" or "20221119" or 2022112* or 2022113* or 202212* or 2023*).dc,dd. | 2079827 |
| 28 | 26 and 27 | 94 |

#### Table S6: Search strategy for Global Health database(s), Global Health 1973 to 2023 Week 38

| **#** | **Searches** | **Results** |
| --- | --- | --- |
| 1 | Human respiratory syncytial virus/ | 7221 |
| 2 | (respiratory syncytial vir* or RSV or hrsv).ti,hw,id. | 7663 |
| 3 | (respiratory syncytial and (rsv or hrsv)).ab. | 5001 |
| 4 | 1 or 2 or 3 [RSV] | 7869 |
| 5 | adults/ | 96854 |
| 6 | (adult? or women or men or man or woman or senior? or elderly or ((old or older) adj2 (patient* or inpatient* or outpatient*))).ti,ab. | 761238 |
| 7 | (adult? or women or men or woman or senior? or elderly or ((old or older) adj2 (patient* or inpatient* or outpatient*))).hw,id. | 509994 |
| 8 | ((aged or ages or year?) adj2 ("18" or "19" or "2#" or "3#" or "4#" or "5#" or "6#" or "7#" or "8#" or "9#")).ti,ab. | 434339 |
| 9 | or/5-8 [adults] | 1035218 |
| 10 | 4 and 9 [RSV + adults] | 1903 |
| 11 | meta-analysis/ or systematic reviews/ | 78419 |
| 12 | ((systematic* adj3 (review* or overview*)) or (methodologic* adj3 (review* or overview*))).ti,ab,hw. | 73003 |
| 13 | ((quantitative adj3 (review* or overview* or synthes*)) or (research adj3 (integrati* or overview*))).ti,ab,hw. | 2776 |
| 14 | (meta analy* or metanaly*).ti,ab,hw,id. | 56365 |
| 15 | (cochrane or evidence report).jx. | 1074 |
| 16 | (comparative adj3 (efficacy or effectiveness)).ti,ab,hw. | 2339 |
| 17 | or/11-16 [adapted from "SR / MA / HTA / ITC - MEDLINE, Embase, PsycInfo. In: CADTH Search Filters Database. Ottawa: CADTH; 2022: <https://searchfilters.cadth.ca/link/33>." Accessed 2022-10-28.] | 97749 |
| 18 | 10 and 17 | 67 |
| 19 | limit 18 to yr="1995-current" | 67 |
| 20 | risk factor/ or health inequalities/ | 354439 |
| 21 | (determinant* or risk or risks or probabilit* or probable or susceptib* or vulnerab* or inequalit* or inequit* or likely or predisposed or likelihood).ti. | 260756 |
| 22 | (determinant* or risk or risks or probabilit* or probable or susceptib* or vulnerab* or inequalit* or inequit* or likely or predisposed or likelihood).ab. /freq=2 | 540447 |
| 23 | or/20-22 | 731070 |
| 24 | 10 and 23 | 418 |
| 25 | editorial.pt. | 3033 |
| 26 | case report?.ti. | 19765 |
| 27 | 24 not (25 or 26) | 418 |
| 28 | limit 27 to yr=2022-current | 67 |

#### Search strategy for ProQuest Public Health limited to systematic review: 1 result

(((TITLE(respiratory syncytial vir* or RSV or hrsv) OR SUBJECT(respiratory syncytial vir* or RSV or hrsv) OR MJMESH( Respiratory Syncytial Virus Infections)) OR ABSTRACT(respiratory syncytial AND (rsv OR hrsv))) AND (MESH(adult) OR TITLE(adult or adults or women or men or man or woman or senior or seniors or elderly or ((old or older) NEAR/2 (patient* or inpatient* or outpatient*))) OR ABSTRACT(adult or adults or women or men or man or woman or senior or seniors or elderly or ((old or older) NEAR/2 (patient* or inpatient* or outpatient*))))) AND (MESH(meta-analysis) or MESH(systematic review) or MESH(meta-analysis as topic) or MESH(systematic review as topic) OR TITLE((systematic* NEAR/3 (review* or overview*)) or (methodologic* NEAR/3 (review* or overview*))) OR ABSTRACT((systematic* NEAR/3 (review* or overview*)) or (methodologic* NEAR/3 (review* or overview*))) OR TITLE((quantitative NEAR/3 (review* or overview* or synthes*)) or (research NEAR/3 (integrati* or overview*))) OR ABSTRACT((quantitative NEAR/3 (review* or overview* or synthes*)) or (research NEAR/3 (integrati* or overview*))) OR TITLE(meta analy* or metanaly*) OR ABSTRACT(meta analy* or metanaly*) OR TITLE(comparative NEAR/3 (efficacy or effectiveness)) OR ABSTRACT(comparative NEAR/3 (efficacy or effectiveness))) AND pd(20221113-20230928)

#### Search strategy for ProQuest Public Health: 35 results

(((TITLE(respiratory syncytial vir* or RSV or hrsv) OR SUBJECT(respiratory syncytial vir* or RSV or hrsv) OR MJMESH( Respiratory Syncytial Virus Infections)) OR ABSTRACT(respiratory syncytial AND (rsv OR hrsv))) AND (MESH(adult) OR TITLE(adult or adults or women or men or man or woman or senior or seniors or elderly or ((old or older) NEAR/2 (patient* or inpatient* or outpatient*))) OR ABSTRACT(adult or adults or women or men or man or woman or senior or seniors or elderly or ((old or older) NEAR/2 (patient* or inpatient* or outpatient*))))) AND (MESH(risk factor) OR MESH(Social Determinants of Health) OR MESH(Health Inequities) OR TITLE(determinant* or risk or risks or probabilit* or probable or susceptib* or vulnerab* or inequalit* or inequit* or likely or predisposed or likelihood) OR ABSTRACT(determinant* or risk or risks or probabilit* or probable or susceptib* or vulnerab* or inequalit* or inequit* or likely or predisposed or likelihood)) AND pd(19950101-20231231)

#### Table S7: Study Inclusion and Exclusion Criteria

|  | **Inclusion** | **Exclusion** |
| --- | --- | --- |
| **Population** | - Adults, including but not restricted to individuals 50 years of age and older and individuals 18 years of age and older with underlying medical conditions. | - Infant and children |
| **Intervention** | NA | NA |
| **Control** | NA | NA |
| **Outcome** | - Medically attended RSV respiratory tract infection (e.g., outpatient and/or ambulatory visits) - Hospitalization associated with RSV infection - ICU admission associated with RSV infection - Death associated with RSV infection | - Outcome not associated with RSV infection |
| **Study design** | - Systematic review and/or meta-analysis - Primary evidence studies (e.g., experimental, quasi-experimental and non-experimental studies) | - Narrative reviews - Guidelines - Editorials, commentary - Conference abstract - Case report and case series |

ICU; intensive care unit, NA; not applicable, RSV; respiratory syncytial virus.

#### Table S8: Risk Factors of Interest and Associated ICD-10-CA Codes

| **Risk factor/comorbidity** | **ICD-10-CA Codes** |
| --- | --- |
| Respiratory tract infection | J09-J22 excluding J12.1, J20.5 and J21.0 |
| Chronic obstructive pulmonary disease | J44 |
| Immunocompromising conditions | B20, B59, B97.3, D70-D73, D76, D80-D84, D89, M05-M08, M30-M34, M35.0, M35.9, Q89.0, Z21, Z48.2, Z51.0, Z51.1,Z94, C00-C26, C30-C34, C37-C41, C43-C58, C60-C86, C88, C90-C96, D03, D46, Z85 |
| Cardiovascular Disease | Myocardial infarction: I21, I22, I25.2  Congestive Heart Failure: I11.0, I13.0, I13.2, I25.5, I42.0, I42.5, I42.6, I42.7, I42.8, I42.9, I43, I50, P29.0  Peripheral Vascular Disease: I70, I71, I73.1, I73.8, I73.9, I77.1, I79.0, I79.1, I79.8, K55.1, K55.8, K55.9, Z95.8, Z95.9.  Cerebrovascular Disease: G45, G46, H34.0, H34.1, H34.2, I60, I61, I62, I63, I64, I65, I66, I67, I68 |
| Diabetes | E10-E14 |
| Chronic Kidney Disease | N18 |
